# The Role of Genetic Testing in Diagnosis and Care of Inherited Cardiac Conditions in a Specialised Multidisciplinary Clinic

**DOI:** 10.1101/2022.02.04.22270485

**Authors:** Fergus Stafford, Neesha Krishnan, Ebony Richardson, Alexandra Butters, Sophie Hespe, Charlotte Burns, Belinda Gray, Caroline Medi, Natalie Nowak, Julia C Isbister, Hariharan Raju, David Richmond, Mark P Ryan, Emma S Singer, Raymond W Sy, Laura Yeates, Richard D Bagnall, Christopher Semsarian, Jodie Ingles

**Affiliations:** Cardio Genomics Program at Centenary Institute, The University of Sydney, Sydney, Australia; Centre for Population Genomics, Garvan Institute of Medical Research, and UNSW Sydney, Sydney, Australia; Centre for Population Genomics, Murdoch Children’s Research Institute, Melbourne, Australia; Faculty of Medicine and Health, The University of Sydney, Sydney, Australia; Agnes Ginges Centre for Molecular Cardiology at Centenary Institute, The University of Sydney, Sydney, Australia; Department of Cardiology, Royal Prince Alfred Hospital, Sydney, Australia; Faculty of Medicine, Health and Human Sciences, Macquarie University, Sydney, Australia

**Keywords:** Inherited cardiac conditions, sudden cardiac death, genetic testing

## Abstract

**Background:** The diagnostic yield of genetic testing for inherited cardiac diseases is up to 40% and primarily indicated for screening of at-risk relatives. Here we evaluate the role of genomics in diagnosis and management among consecutive individuals attending a specialised clinic and identify those with highest likelihood of having a monogenic disease.

**Methods:** Retrospective audit of 1697 consecutive, unrelated probands referred to a specialised, multidisciplinary clinic between 2002 and 2020. A concordant clinical and genetic diagnosis was considered solved. Cases were classified as likely monogenic based on a score comprising a positive family history, young age at onset and severe phenotype, whereas low scoring cases were considered to have a likely complex aetiology. The impact of a genetic diagnosis was evaluated.

**Results:** A total of 888 probands fulfilled inclusion criteria, and genetic testing identified likely pathogenic or pathogenic (LP/P) variants in 330 individuals (37%), and suspicious variants of uncertain significance (VUS) in 73 (8%). Research-focused efforts identified 46 (5%) variants, missed by conventional genetic testing. Where a variant was identified, this changed or clarified the final diagnosis in a clinically useful way for 51 (13%). The yield of suspicious VUS across ancestry groups ranged from 15-20%, compared to only 10% among Europeans. Even when the clinical diagnosis was uncertain, those with the most monogenic disease features had the greatest diagnostic yield from genetic testing.

**Conclusion:** Research-focused efforts can increase the diagnostic yield by up to 5%. Where a variant is identified, this will have clinical utility beyond family screening in 13%. We demonstrate the value of genomics in reaching an overall diagnosis, and highlight inequities based on ancestry. Acknowledging our incomplete understanding of disease phenotypes, we propose a framework for prioritising likely monogenic cases to solve their underlying cause of disease.

## INTRODUCTION

Inherited cardiac conditions are associated with significant morbidity and mortality and can result in sudden cardiac death (SCD) at a young age. Clinical genetic testing is a key aspect of clinical management (1), currently used primarily for cascade testing of asymptomatic at-risk relatives. For some, a genetic result can also change diagnosis, clarify prognosis, and guide clinical management. However, the diagnostic yield of cardiac genetic testing remains low, with only ∼40% having a likely pathogenic or pathogenic variant (LP/P) identified (2,3). Critical gaps remain in our understanding of the phenotypic spectrum and genetic architecture of these diseases, including growing acknowledgement of a role for complex polygenic contributions to disease (4-6).

Efforts to improve diagnostic yield of genetic testing thus far have largely focused on patients who fulfil clinical diagnostic criteria for an inherited cardiac condition. It is unclear what proportion have undifferentiated cardiac pathology that does not meet widely-accepted specific clinical diagnostic criteria, i.e. undiagnosed cardiac disease. The role genetic testing can play in this setting is far less clear, but presents an exciting opportunity to improve the diagnostic yield through gene discovery and mechanistic insights, particularly among those with features suggestive of a monogenic cardiac disease.

Individuals with monogenic diseases are more likely to have a younger age at onset (8,9) and a positive family history, compared to their gene negative counterparts (4,10). Recent studies have shown HCM patients with causative variants in sarcomere genes have more severe disease and worse outcomes compared to sarcomere-negative patients (4,11), and more generally individuals with phenotypic extremes are enriched for rare, large effect size variants (12,13). Determining the likelihood of monogenic disease could provide a powerful tool in discriminating between monogenic and complex disease etiology, and allow prioritisation of cases where additional gene discovery efforts will be of greatest utility.

Here we describe our experience of evaluation and diagnosis of individuals seen in a specialised multidisciplinary cardiac genetic clinic over an 18-year period. Those with a concordant clinical and genetic diagnosis were considered solved, and we report the proportion in which the final diagnosis was clarified or changed due to genetic testing. We describe the role of genomics in elucidating an overall diagnosis and propose a simple scoring system for prioritising difficult unsolved cases most likely to have a monogenic disease basis. Clarifying the underlying diagnosis in these unsolved cases may provide important insights about disease mechanisms and lead to increased yields of cardiac genetic testing with potential for important clinical and prognostic implications worldwide.

## METHODS

### Study population

Consecutive, unrelated probands referred to a specialised, multidisciplinary clinic at Royal Prince Alfred Hospital (Sydney, Australia) between 2002 and 2020 were evaluated. These clinics were initially established for management of hypertrophic cardiomyopathy (HCM); however, since 2003 have expanded to include inherited cardiomyopathies, arrhythmia syndromes and sudden cardiac death evaluation (14). Probands were included if they, or at least one family member, were: seen in the clinic, had definite or probable cardiac disease, and underwent comprehensive clinical investigation and genetic testing. Families where the only affected family member was deceased and not evaluated during life were excluded. Ethics approval was granted by Sydney Local Health District Human Research Ethics Committee (X20-0522 & 2020/ETH03142).

### Clinical investigations and baseline diagnosis

#### Cardiac evaluation

Individuals were evaluated per clinical recommendations at the time of consultation. Most patients underwent transthoracic echocardiography and 12-lead electrocardiography (ECG) in clinic as a minimum. Where this was not possible, results of previous investigations were requested from the referring clinician or hospital. Further investigations were conducted at the discretion of the treating cardiologist, and included: modified “high” right precordial ECG (V1 and V2 cranially-displaced into 2nd and 3rd intercostal spaces); 24-hour ambulatory Holter monitoring (including 12-lead and/or modified high right precordial lead positions); exercise stress testing; signal-averaged ECG; and drug provocation tests (including flecainide or ajmaline challenge depending on the year performed). Cardiac magnetic resonance (CMR) imaging was routinely performed to identify structural changes including myocardial noncompaction where echocardiography was negative or inconclusive, and when assessing for arrhythmogenic cardiomyopathy (ACM) (15). Cardiac investigations were reported by cardiologists as per clinical practice and according to accepted guidelines (16-18).

*Family history* was evaluated by experienced cardiac genetic counsellors from 2003 and included a 3-generation pedigree identifying affected relatives and any significant cardiac events such as a SCD, cardiac transplant etc. Details were confirmed with medical records and postmortem reports where available.

#### Baseline phenotype and clinical diagnosis

The clinical diagnosis after baseline evaluation was classified as definite or uncertain. A definite clinical diagnosis included any individual meeting diagnostic criteria, with or without atypical features. An uncertain clinical diagnosis included those with an abnormal but uncertain phenotype, such as possible (borderline) diagnoses and undiagnosed (unclassified) cardiac disease. Cases were adjudicated to exclude those with likely physiological changes, and discussed in regular multidisciplinary meetings with other clinical team members as needed.

#### Time to diagnosis

For those solved using additional genetic testing research efforts, time to diagnosis in years from the first presentation in the clinic to a genetic diagnosis was ascertained. Time to diagnosis was calculated within two decades: 2002-10 and 2011-20.

### Genetic testing and variant interpretation

Cardiac genetic testing including targeted gene panels, whole exome and whole genome testing was performed clinically or on a research basis, and as per best practice at the time of sequencing. Variants were assessed and classified using *MYH7*-modified American College of Medical Genetics and Genomics and Association of Molecular Pathologists (ACMG/AMP) standards for sequence variant interpretation (19). Likely pathogenic and pathogenic (LP/P) variants were considered causative/solved. Where no variants were identified, or were classified as benign, likely benign, or non-suspicious VUS, genetic testing was considered to be negative, i.e. unsolved. Patients with variants of uncertain significance (VUS) deemed suspicious but lacking critical evidence were not considered unsolved and are shown separately. Suspicious VUS were absent or very rare, i.e. <0.004% for hypertrophic cardiomyopathy (HCM) and more stringent for other less prevalent diseases (19,20), in population databases (Genome Aggregation Database; gnomAD);(21) occurred in a gene with definitive gene-disease association relevant to the baseline phenotype, and predicted to alter the protein sequence. Genes with robust gene-disease association at time of interpretation were considered Tier 1 genes, while Tier 2 genes were those with less well characterised clinical validity (Supplementary Table 1) (22-25). Mitochondrial DNA variants were not routinely evaluated, except for those who underwent whole genome sequencing as part of gene discovery studies.

### Likelihood of monogenic disease

Based on the available literature and our experiences in the clinical setting, we identified a severe phenotype (4,11,13), young age of disease onset (8,9) and positive family history (10) as characteristics more likely to be associated with monogenic disease, as opposed to complex disease arising from environmental factors and/or the accumulation of common genetic variants with small effect sizes. The scoring system and disease specific high risk/severe phenotype definitions are shown in Figure 1A, incorporating characteristics from the family unit. A higher score indicated a greater likelihood of monogenic disease.

**Figure 1:**
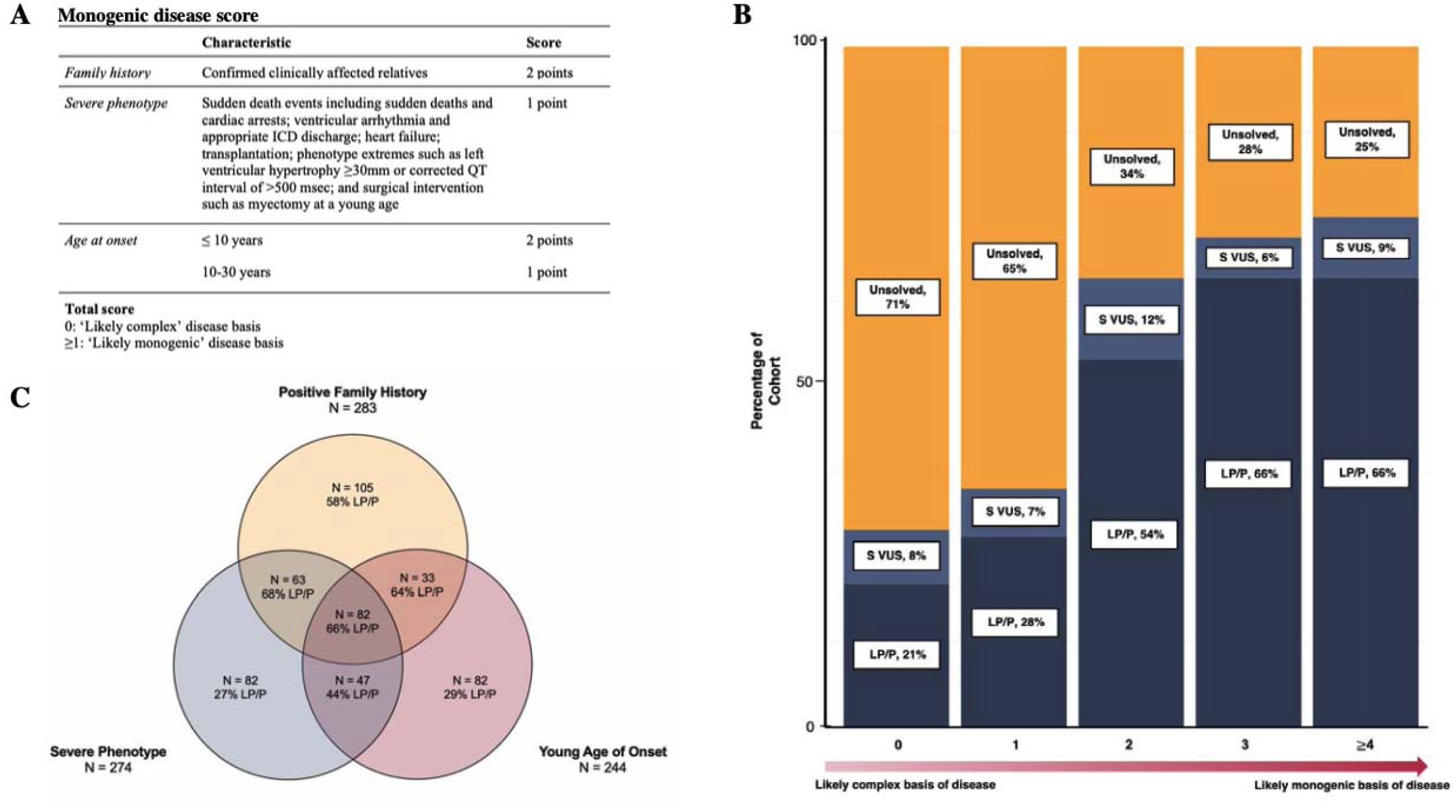
(A) Monogenic disease score, (B) Diagnostic yield of genetic testing based on monogenic disease scores, (C) Frequency and diagnostic yield among individuals with different clinical features suggesting monogenic disease. Abbreviations: LP/P, likely pathogenic and pathogenic; S-VUS, suspicious variant of uncertain significance; ICD, implantable cardioverter defibrillator.

### Final diagnosis and clinical impact

Solved cases were those where the clinical diagnosis and genetic findings were concordant and adequately explained disease. When additional research was required to make a genetic diagnosis, these were considered ‘solved on review’. Cases were considered ‘unsolved’ if no genetic cause for their disease was identified. If no genetic variants were identified with one or more features indicative of monogenic disease present, those individuals were considered unsolved and more likely to have a monogenic basis (‘likely monogenic’). Conversely, individuals with no causative genetic variant and without these features were considered unsolved and more likely to have complex aetiology (‘likely complex’).

Among solved cases, we evaluated whether the genetic result clarified inheritance risk, family screening recommendations, frequency and type of cardiac investigations, additional specialist consultations (e.g. neurological review), changes to medication, and lifestyle advice arising from their genetic diagnosis.

### Ancestry

Self-reported ancestry was recorded for most probands and classified according to the Australian Standard Classification of Cultural and Ethnic Groups (26). Ancestry groups included European, East Asian, South and Central Asian, Middle-Eastern and North African or other ancestry (including less frequent ancestries such as Sub-Saharan African, Oceanian and People of the Americas). We compared the proportion of patients with LP/P variants, suspicious VUS, and unsolved between all groups to assess diagnostic yield. Representation of ancestry groups was compared to the Greater Sydney population using 2016 Census data (27). Census data include a proportion who identify as ‘Australian’, who have typically included a high number of Anglo-Celtic individuals, and were considered to have European ancestry (28).

### Statistical analysis

Data were analysed using R (Version 4.1.0) and GraphPad Prism (Version 9.2.0). Descriptive statistics were performed, including chi-squared tests for categorical variables and student’s t-test for continuous variables. A p-value of <0.05 was considered statistically significant. R packages *ggplot2* and *ggalluvial* were used for figures.

## RESULTS

### Study population and baseline clinical diagnoses

There were 1697 probands who attended the specialised clinic between 2002 and 2020, and 888 met inclusion criteria, while 809 were excluded due to no genetic testing performed, no living affected family member seen in the clinic, cardiac findings determined to be physiological, or limited information or clinical evaluation performed (Figure 2). Of 888, a genetic cause of disease was identified in 403 (45%), while 485 (55%) remained unsolved.

**Figure 2:**
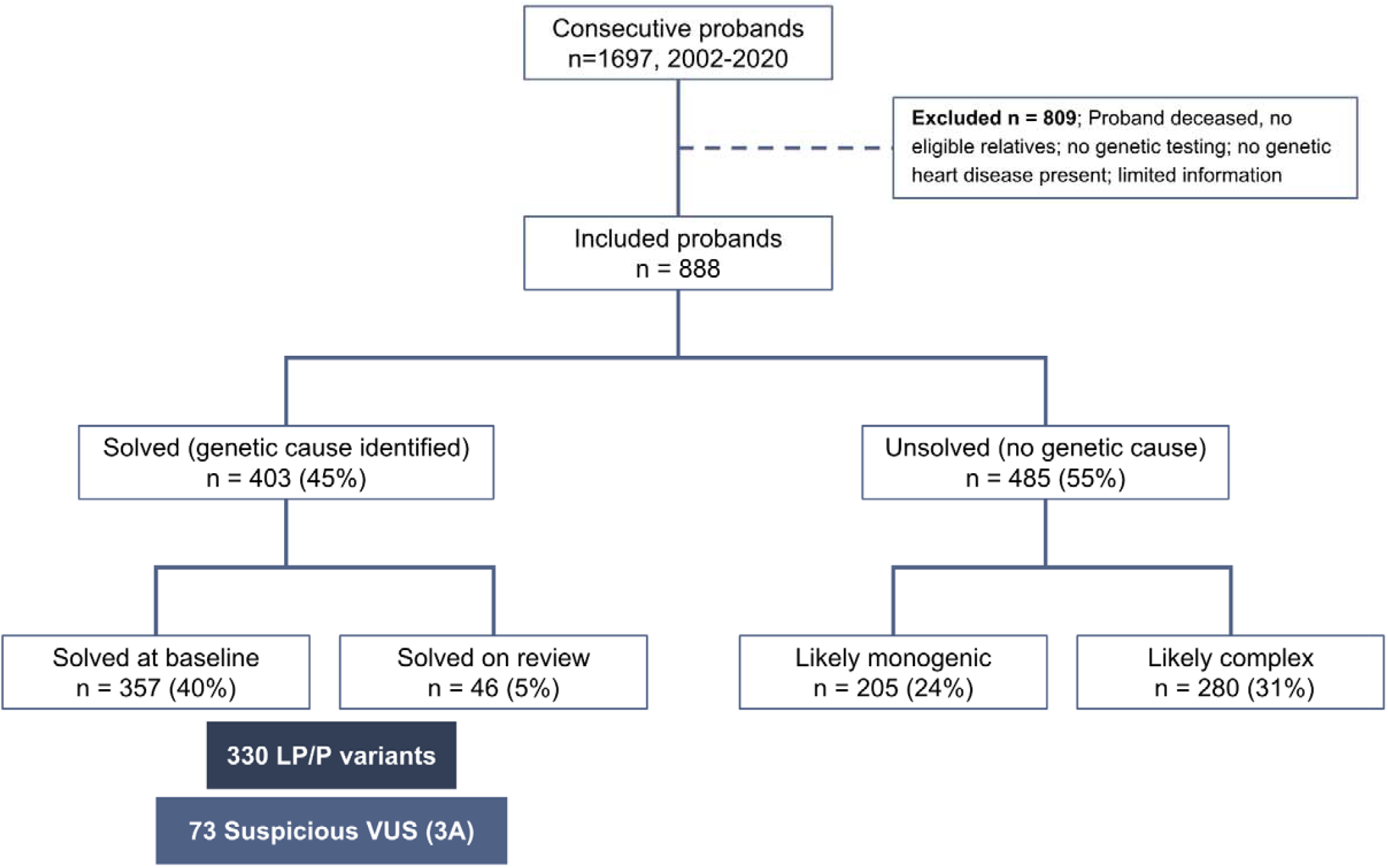
Flow diagram of included probands and their outcomes. Abbreviations: LP/P, likely pathogenic and pathogenic variants; VUS, suspicious variant of uncertain significance.

The demographic and clinical characteristics are summarised in Table 1. Following baseline evaluation, a definite clinical diagnosis was made in 762 (86%) of the total cohort. The remaining 126 (14%) had uncertain clinical diagnoses, including 21 (17%; 2% of the total audit cohort) with undiagnosed primary cardiac phenotypes, 36 (28%) with unexplained resuscitated sudden cardiac arrest (SCA), and 69 (55%) with possible disease. The majority of those with a definite clinical diagnosis at baseline were diagnosed with HCM (589/762, 77%), Brugada syndrome (n=50; 7%), or long QT syndrome (n=44; 6%). Less common diagnoses, accounting for 1-5% of those with a definite diagnosis, included dilated cardiomyopathy (DCM), left-ventricular noncompaction (LVNC), arrhythmogenic cardiomyopathy (ACM), and catecholaminergic polymorphic ventricular tachycardia (CPVT). Noonan syndrome, restrictive cardiomyopathy, and sick sinus syndrome each accounted for <1%.

**TABLE 1:** Clinical characteristics of included probands.

|  | **Definite diagnosis** | **Uncertain clinical diagnosis** | **Total cohort** |
| --- | --- | --- | --- |
| Age, years | 56.0 ± 16.5 | 46.1 ± 15.9 | 54.8 ± 16.7 |
| Age at diagnosis, years | 42.3 ± 16.6 | 36.5 ± 16.1 | 41.6 ± 16.6 |
| Male sex | 487 (64) | 71 (56) | 558 (63) |
| Severe phenotype | 209 (27) | 65 (52) | 274 (31) |
| Sudden cardiac events | 84 (11) | 51 | 135 |
| Heart failure | 7 (<1) | 0 (0) | 7 (<1) |
| Transplantation | 9 (1) | 3 (<1) | 12 (14) |
| Phenotype extremes | 74 (10) | 8 (6) | 82 (9) |
| Ventricular arrhythmia | 30 (4) | 3 (2) | 33 (4) |
| Positive family history | 247 (32) | 36 (29) | 283 (32) |
| Age at diagnosis ≤30 years | 192 (25) | 52 (41) | 244 (27) |
| Age at diagnosis ≤10 years | 7 (< 1) | 0 (0) | 7 (<1) |
| Inherited cardiomyopathy | 662 (87) | 62 (49) | 724 (82) |
| Inherited arrhythmia syndrome | 100 (13) | 24 (19) | 124 (14) |
| Undiagnosed | 0 (0) | 40 (32) | 40 (5) |
| Total | 762 (86) | 126 (14) | 888 (100) |

Data are shown as mean ± standard deviation, or n (%).

### Genetic findings

Genetic testing identified LP/P variants in 330 (37%) of the total cohort, suspicious VUS in 73 (8%), and the remainder had no variants of interest (n=485; 55%; Figure 3A). There were 209 distinct variants reported and classified as LP/P (Supplementary Table 2) and 73 as suspicious VUS (Supplementary Table 3). The yield of LP/P variants and suspicious VUS were similar for those with inherited cardiomyopathies and inherited arrhythmia syndromes (Figure 3B).

**Figure 3:**
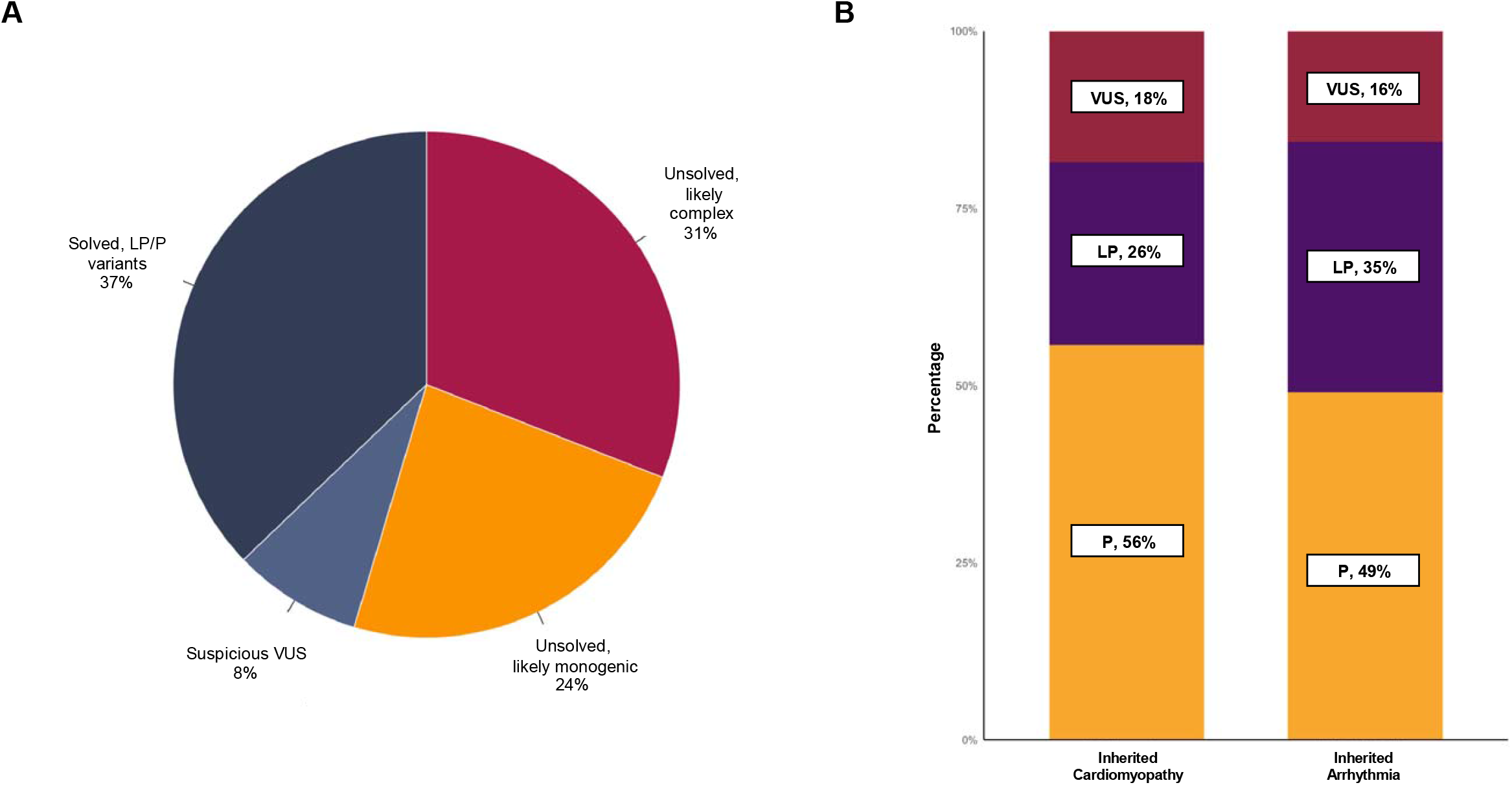
(A) Diagnostic yield of genetic testing among all patients (B) Proportion of solved cases with variants classified as pathogenic, likely pathogenic, or suspicious VUS. Abbreviations: P, pathogenic; LP, likely-pathogenic; S VUS, suspicious variant of uncertain significance.

Variants classified as suspicious VUS (n=73) were most commonly identified in *MYBPC3* (17/73, 23%), followed by *MYH7* (n=15, 21%), *TNNI3* (n=6, 8%), *TNNT2* (n=5, 7%), *KCNH2* (n=4, 5%), *FLNC* (n=4, 5%) and *PRKAG2* (n=3, 4%). While these variants lacked critical evidence supporting pathogenicity, existing evidence included rarity in the general population (PM2; n=73, 100%), *in silico* tools supporting a deleterious effect (PP3; n=57, 78%); enrichment in cases compared to controls as a supporting (PS4_supporting; n=17, 23%), or moderate (PS4_moderate; n=5, 7%) level of evidence; whether the variant was located in a hotspot region (PM1; n=5; 7%; and PM1_supporting n=2, 3%); segregation with disease in a family (PP1, cosegregation with greater than three meioses; n=4; 6%). Other criteria applied included PM6 (assumed *de novo* without confirmation of paternity and maternity), PVS1_moderate (null variant in a gene where loss of function is a known disease mechanism), PS3_supporting (well-established functional studies), applied once each.

### Achieving a final genetic diagnosis

There were 357 (40%) of the total cohort who were considered solved using standard clinical-genetic approaches (solved at baseline), and an additional 46 (5%) solved on review following additional effort. Evaluation of tier-2 gene lists (via whole exome and genome sequencing) in individuals not solved at baseline identified LP/P variants in a further 17/370 (5%) and suspicious VUS in 7 (2%).

A genetic diagnosis clarified the clinical diagnosis for 51 (13%) of the 403 patients with an LP/P variant. (Figure 4). This included six individuals with an undiagnosed primary cardiac disease in whom genetic testing provided a diagnosis, including a *TTN* truncating variant in a patient with conscious VT, mitral valve prolapse and mild LV dysfunction; an *NDUFB11* variant in two individuals with atypical LVH, clarifying the phenotype as histiocytoid cardiomyopathy; a missense variant in *CACNA1C* in one individual with an atypical LVH and arrhythmia phenotype; a missense *DES* variant in one individual with complete infranodal heart block and subsequent VF arrest; and an *SCN5A* truncating variant in one individual with atrial flutter and complete heart block.

**Figure 4:**
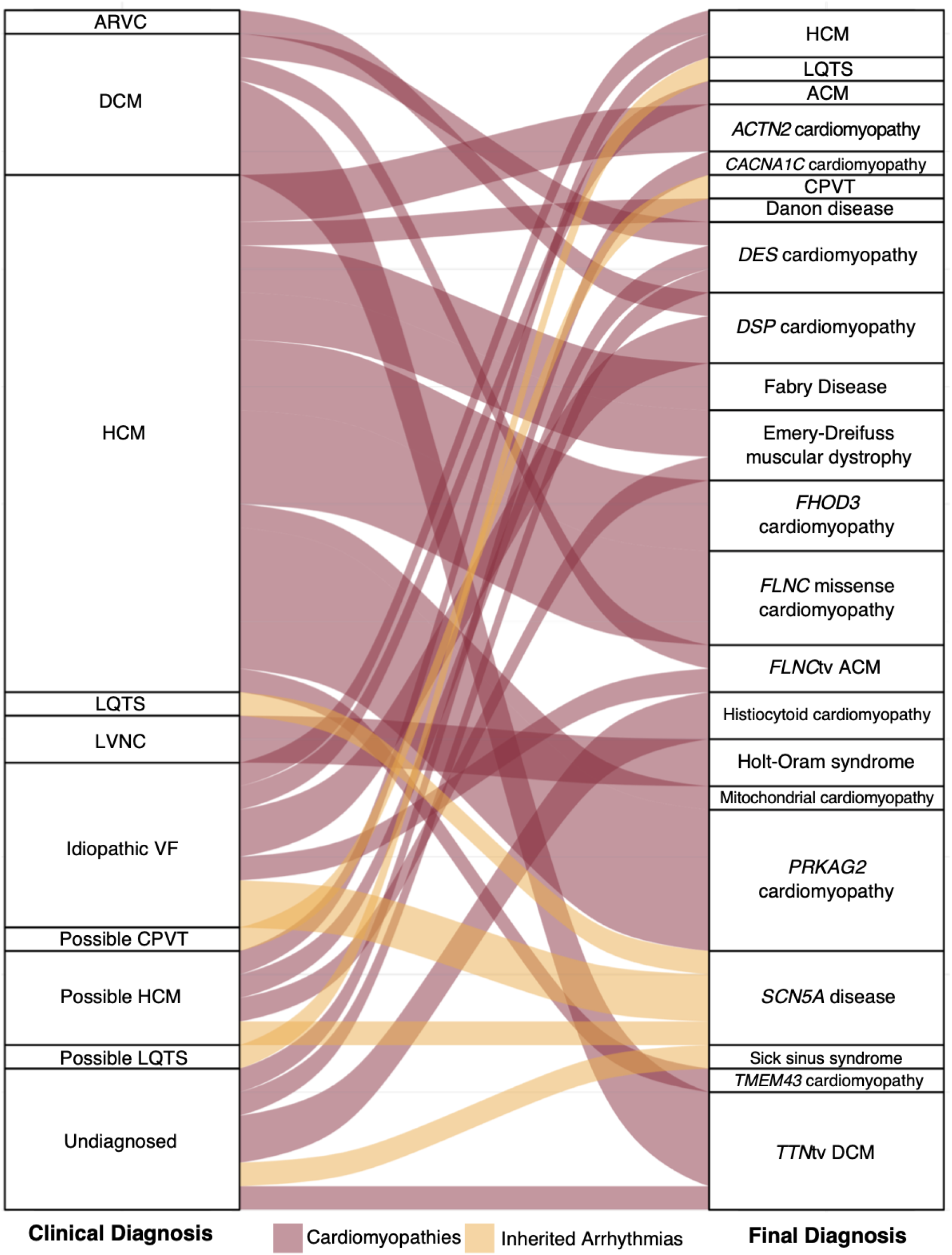
Impact of genomics in 51 patients where genetic testing clarified the overall diagnosis. Baseline phenotype of ARVC included those who met 2010 Task Force Criteria. Abbreviations: ARVC, arrhythmogenic right ventricular cardiomyopathy; DCM, dilated cardiomyopathy; HCM, hypertrophic cardiomyopathy; LQTS, long QT syndrome; LVNC, left-ventricular non-compaction; CPVT, catecholaminergic polymorphic ventricular tachycardia; ACM, arrhythmogenic cardiomyopathy.

Among the 46 (5%) solved on review following additional effort, a genetic diagnosis was most commonly achieved using research-focused genetic testing (Figure 5). Often multiple factors were required to achieve the final diagnosis. Time-to-diagnosis in this group was prolonged for those clinically diagnosed in the years 2002-2010 (n=16, mean 11.9 years SD 5.7) compared with 2011-2020 (n=30, mean 2.7 years SD 5.4; p<0.01).

**Figure 5:**
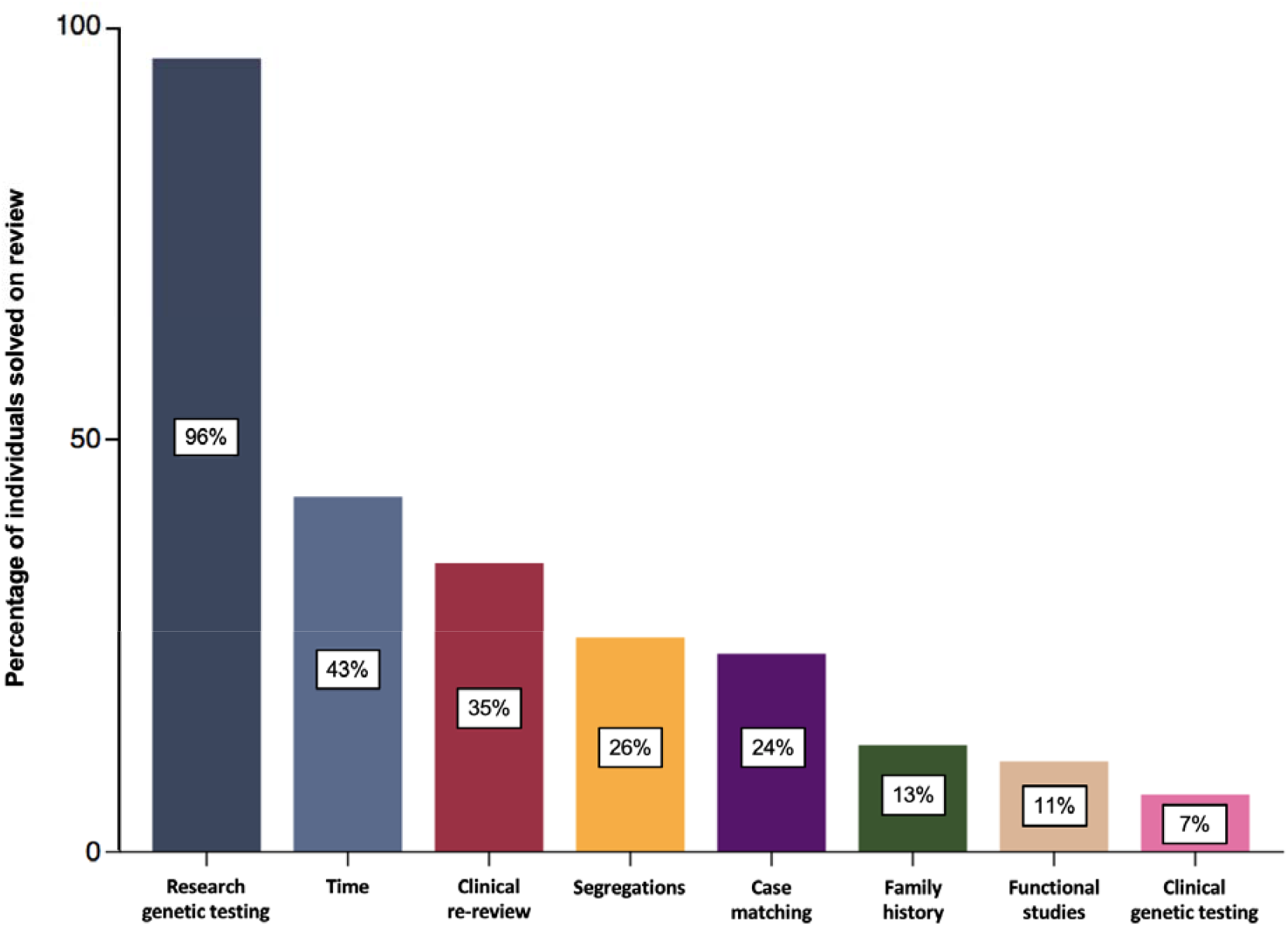
Reasons for achieving a final diagnosis among 46 patients where a genetic diagnosis was made following additional research efforts. The proportion of patients ‘solved on review’ in whom each reason contributed to achieving a diagnosis. Multiple reasons often contributed to an individual’s diagnosis.

Among cases solved on review, 20/46 (43%) had an uncertain baseline clinical diagnosis. The most common phenotype at baseline evaluation was HCM (20; 43%) or possible HCM (borderline LVH; 6, 13%). All had undergone prior genetic testing that failed to identify a cause, with reasons including genes not previously tested, variants located in deep intronic regions, or lack of available evidence for pathogenicity at the time of testing (e.g. reported proband counts, segregations). Subsequent sequencing identified six with *MYBPC3* variants (4 deep intronic splice-site altering, 1 synonymous affecting splicing and 1 missense variant(29)); four with missense variants in *FLNC*; three with truncating or canonical splice-site variants in *FHOD3* (30); two with missense variants in *ACTN2* (31); two with missense variants in *MYH7*; one each with a missense variant in *TNNI3, PRKAG2, MT-TI*; a *de novo* variant in *DES*; one individual with asymmetric LVH and high burden of ventricular ectopy with the pathogenic p.S358L variant in *TMEM43*; and one with SCD <30 years and postmortem findings suggesting possible HCM but family history of Brugada syndrome and sinus node disease with a balanced translocation impacting *SCN5A* (32). Two individuals presented with LVNC, and genetic testing identified putative loss of function variants in *TBX5* with subsequent clinical review confirming a diagnosis of Holt-Oram syndrome (33).

### Ancestry

Self-reported ancestry was available for 541/608 probands (89%). The majority were of European ancestry (n=403, 75%), 40 (8%) East-Asian, 30 (6%) South Asian, 43 (8%) Middle-Eastern and North African, and 26 (5%) categorised as other (comprised Oceanian, Sub-Saharan African, and peoples of the Americas’ ancestries).

The diagnostic yield differed based on ancestry group (Figure 6). Middle-Eastern and North African patients had the lowest yield of LP/P variants (12/43, 28%), a high rate of suspicious VUS (n=8, 19%), with more than half of patients having no clear genetic cause for their disease (n=23, 54%). Conversely, the diagnostic yield was the highest among probands with South Asian (LP/P, 67%), European (LP/P, 57%) and other (LP/P, 62%) ancestry groups. When compared to Europeans (41/403, 10%), the yield of suspicious VUS was higher in other ancestry groups (15-20%; chi-squared test, p<0.05).

**Figure 6:**
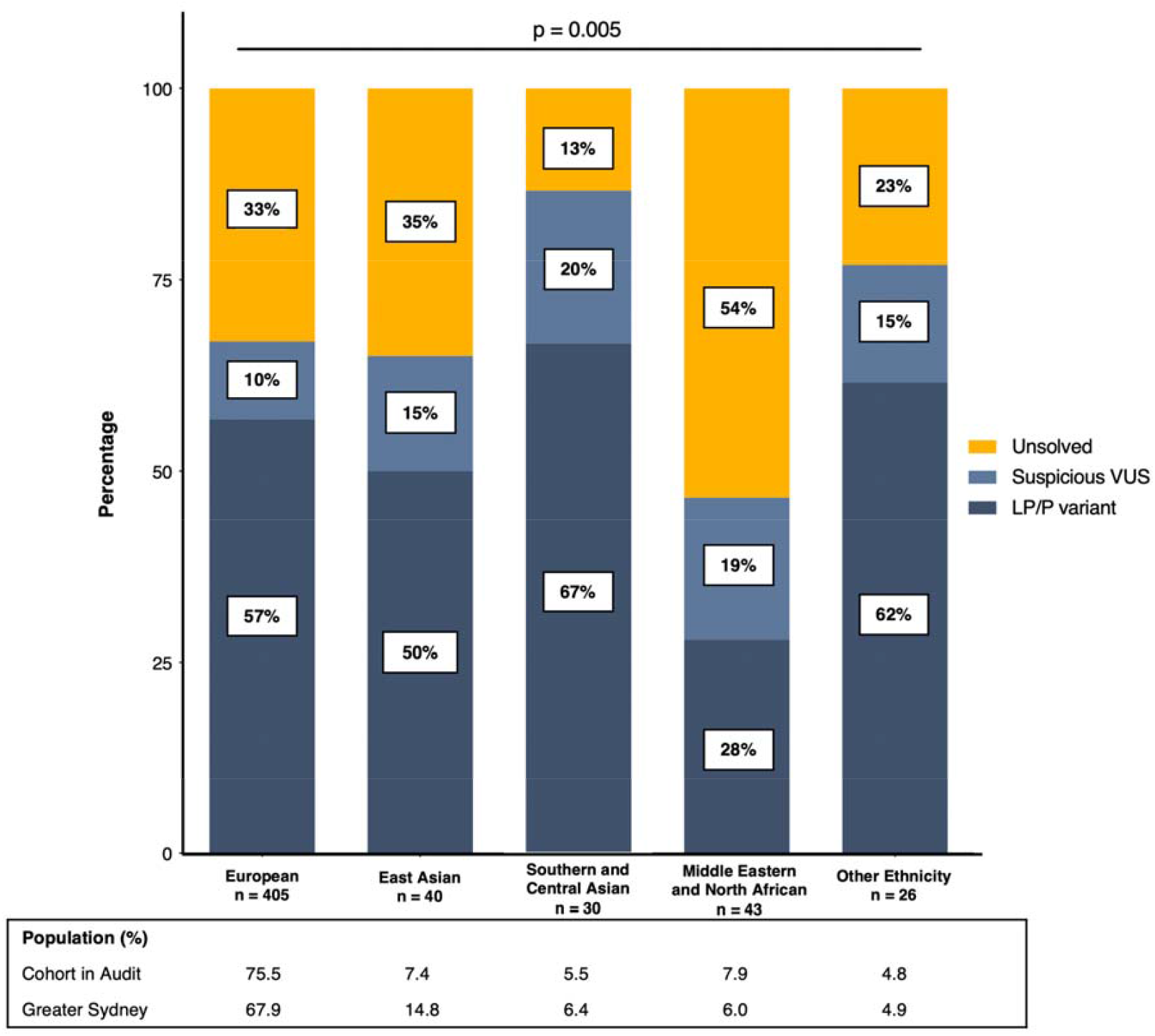
Diagnostic yield of genetic testing by major ancestry groups. ‘Other ancestry’ includes Oceanian, People of the Americas. Proportion of suspicious VUS compared between ancestry groups using chi-square. Abbreviations: LP/P, likely pathogenic and pathogenic variants; VUS, variant of uncertain significance.

Comparison with the prevalence of ancestry groups in the clinic cohort versus Greater Sydney was as follows: East Asian (7.4% versus 14.8%), South Asian (5.5% versus 6.4%), Middle Eastern and North African (7.9% versus 6.0%) and European patients (75.5% versus 67.9%).

### Impact of a genetic diagnosis

Among 26 of 46 (57%) who were solved on review, the genetic diagnosis resulted in a change in management and risk stratification for the proband and/or their family. There was a change in the inheritance risk in 11 (22%), including eight where X-linked inheritance was confirmed, two with *de novo* variants, and one with a mitochondrial variant that confirmed maternal inheritance. Clinical screening recommendations for family members were altered in 14 (30%), such as the need for CMR imaging, drug provocation challenge, and/or referral to other (non-cardiac) specialised clinics. Additional specialist review was indicated for 16 (35%) probands, including neurologist for three (7%) with Emery-Dreifuss muscular dystrophy due to *FHL1* variants previously diagnosed as HCM, three (7%) with myofibrillar myopathy due to *DES* variants, and two (4%) undiagnosed individuals with histiocytoid cardiomyopathy due to variants in *NDUFB11*; clinical geneticist (n=5, 11%); and referral to a specialised Fabry disease clinic (n=2, 4%) including one who subsequently commenced enzyme-replacement therapy. One patient found to have Danon disease was expedited for review and subsequent cardiac transplantation (34).

### Monogenic disease score

There were 283 (32%) of the total cohort with a positive family history of disease, 274 (31%) were considered to have a severe phenotype and 244 (27%) diagnosed 30 years of age including 7 (0.8%) diagnosed 10 years of age. These characteristics were used to calculate their likelihood of having monogenic disease. Most individuals accumulated few points, with 394 (45%) scoring 0, 162 (18%) scoring 1, 153 (17%) scoring 2, 97 (11%) scoring 3, and 82 (9%) with scores 4. The diagnostic yield increased for those with higher scores, i.e. increasing likelihood of monogenic disease (Table 2). The yield of LP/P variants ranged from 21% (those with a score of 0) up to 66% (scores 4; Figure 1A).

**TABLE 2:** Diagnostic yield of genetic testing.

|  | Monogenic disease score |  |  |  |  |
| --- | --- | --- | --- | --- | --- |
|  | 0<br>(likely complex) | 1 | 2 | 3 | ≥4<br>(likely monogenic) |
| *Definite clinical diagnosis* | 362 | 121 | 127 | 85 | 67 |
| Likely pathogenic/pathogenic | 82 (23) | 40 (33) | 77 (61) | 58 (68) | 50 (75) |
| Suspicious VUS | 27 (7) | 9 (7) | 18 (14) | 6 (7) | 6 (9) |
| *Uncertain clinical diagnosis* | 32 | 41 | 26 | 12 | 15 |
| Likely pathogenic/pathogenic | 2 (6) | 7 (17) | 5 (19) | 6 (50) | 4 (27) |
| Suspicious VUS | 3 (9) | 3 (7) | 0 (0) | 0 (0) | 1 (7) |
| *Total cohort* | 394 | 162 | 153 | 97 | 82 |
| Likely pathogenic/pathogenic | 84 (21) | 46 (28) | 82 (54) | 64 (66) | 54 (66) |
| Suspicious VUS | 30 (8) | 12 (7) | 18 (12) | 6 (6) | 7 (9) |

Data are shown as n (%) of each score group. Abbreviations: VUS, variant of uncertain significance.

Among the 494 individuals with at least one of these characteristics suggestive of monogenic disease, the diagnostic yield of genetic testing was lowest among individuals with only a severe phenotype (LP/P; 22/82, 27%), or only an age of disease onset 30 years (LP/P; 24/82, 29%; Figure 1C), each allocated 1 point under our proposed scoring system. When both of these characteristics were present, the yield increased to 44% (21/47, p<0.05). The yield of LP/P variants among individuals with only a positive family history (allocated 2 points) was 58%, and likewise increased when present in combination with a young age of onset (21/33, 64%), severe phenotype (43/63, 68%), or both (54/82, 66%).

Among those with uncertain clinical diagnoses at baseline evaluation, the overall yield of genetic testing was 24/126 (19%). However, stratifying these cases based on their likelihood of having a monogenic disease was able to improve the genetic yield. For those with a monogenic disease score of 0 (i.e. likely complex disease) there were only 2/32 (6%) individuals with a LP/P variant identified. While those with scores of 3 and 4 had a yield of 6/12 (50%) and 4/15 (27%), respectively (Table 2).

The four highest scoring families where a genetic diagnosis could be made are shown in Table 3, and include individuals with atypical and severe clinical presentations and causative variants identified via research-based whole exome or genome sequencing in three, and comprehensive clinical cardiac panel in one.

**TABLE 3:** Clinical and genetic features of high scoring solved and unsolved families.

| Family Code | Score | Proband | Family History | Genetic Testing |
| --- | --- | --- | --- | --- |
| **Unsolved (ongoing research)** |  |  |  |  |
| VE | 5 | Clinically diagnosed with HCM <10 years with moderate LVH. SCD <30 years, with HCM identified as cause of death at post-mortem. | Second-degree relative with confirmed ARVC. | Negative WES. |
| AMZ | 4 | SCD <30 years with moderate hypertrophy and dilatation, and minor RV fibrosis and fatty infiltration at post-mortem. No clinical diagnosis. | Multiple syncope and RV dilatation in one sibling. Diagnostic type 1 Brugada pattern on ajmaline challenge in multiple relatives. | Negative WGS in proband and negative WES in first, second, and third degree relatives. |
| CFR | 4 | Multiple syncope, episodes of dyspnoea, and 6 second sinus pause on loop recorder onset <30 years. Mild LV dilatation on CMR. No clinical diagnosis. | SUD during sleep in parent age <40 years and one sibling with unclassified sinus node disease and PPM. | Negative gene panel and WES. |
| ABM | 4 | SCA <30 years, structurally normal heart with atypical ECG changes. ICD implanted for secondary prevention. | Children with ECG changes including bradycardia, implanted with PPM. | Negative gene panel and WES. |
| **Solved on review** |  |  |  |  |
| EI | 4 | SCA during pregnancy, apical LVH leading to a clinical diagnosis of HCM (ICD implanted). Later decompensated, progressing to severe heart failure. Final diagnosis of *ACTN2* cardiomyopathy. | Multiple affected relatives with a heterogeneous phenotype ranging from asymptomatic to syncope, heart failure, and SCD. | LP variant in *ACTN2* identified via research based linkage analysis.] |
| RY | 4 | Mild apical LVH and atypical ECG changes, clinically diagnosed with HCM | SCD in sibling <20 years. | Negative gene panel, LP variant in *FHL1* identified on |
|  |  | (borderline). Final diagnosis of *FHL1* cardiomyopathy, with neurological review identifying elbow contractures and mild muscular dystrophy/ |  | WES. |
| TD | 4 | SCD <30 years, diagnosed with possible HCM at postmortem investigation. Final diagnosis of *SCN5A* disease. | Family member experienced syncope, multiple relatives subsequently diagnosed with Brugada syndrome. | Negative gene panel, LP translocation in *SCN5A* on WGS. |
| CPV | 3 | Elite athlete, presented with SCA <30 years, but no clinical diagnosis despite extensive investigation. Final diagnosis *DSP* cardiomyopathy. | No family history of disease. | P truncating variant in *DSP* identified on a comprehensive cardiac panel. |

**Abbreviations:** SCD, sudden cardiac death; HCM, hypertrophic cardiomyopathy; ARVC, arrhythmogenic right ventricular cardiomyopathy; SUD, sudden unexpected death with no cause identified; ECG, electrocardiogram; SCA, sudden cardiac arrest; ICD, implantable cardioverter defibrillator; WES, whole exome sequencing; WGS, whole genome sequencing; RV, right ventricle; CMR, cardiac magnetic resonance; PPM, permanent pacemaker; LVH, left ventricular hypertrophy.

### Unsolved cases

There were 485 (55%) patients of the total cohort who remained genetically unsolved after both clinical and genetic investigations, of whom 205/485 (42%) were considered unsolved with likely monogenic disease and 280 (58%) unsolved with a likely complex etiology (Figure 2).

Individuals without a genetic diagnosis but with clinical features suggestive of monogenic disease (n=205) scored 1 and were classified as being likely monogenic. There were 136/205 (66%) with a definite clinical diagnosis at baseline and 69 (34%) with uncertain clinical diagnoses. There were 62 (30%) who had not undergone expanded genetic testing to date. The highest scoring unsolved cases are described in Table 3 and include individuals with variable clinical presentations, family history and research-based whole exome sequencing so far negative.

Among the 280 considered unsolved with a likely complex disease basis, 27 did not have diagnostic clinical findings at baseline evaluation and were classified as uncertain. No individuals had a severe phenotype, positive family history, and all had age at onset over 30 years. There were 93 individuals who had not undergone expanded genetic testing and 207 (74%) had a diagnosis of HCM.

## DISCUSSION

We report our 18-year experience of genetic testing in a specialised cardiac genetic clinic and demonstrate a role for genetic testing in clarifying overall diagnosis and patient management. While genetic testing is indicated for those with a definite clinical diagnosis, our dynamic understanding of the phenotypic spectrum of these diseases, and the presentation of individuals with undiagnosed or uncertain clinical diagnoses underscores the limitations of this approach. We show that the cumulation of factors indicative of monogenic disease, such as severe phenotype, young age at onset and positive family history, can identify those most likely to achieve a genetic diagnosis. Our findings support genetic testing being offered to all individuals with a confirmed or suspected inherited cardiomyopathy or arrhythmia syndrome, and subsequent triaging of cases with characteristics suggestive of a monogenic disease basis for additional, comprehensive research-based sequencing. At present, there are inequities in genetic yield based on ancestry.

Despite decades of research effort, the diagnostic yield of genetic testing for inherited cardiac conditions has barely increased. More stringent variant classification criteria have raised the threshold required to consider a variant the cause of disease (19,35), and gene curation efforts attempt to guide consideration of variants only in genes with robust clinical validity for the phenotype in question (22,23,25,36). It is widely accepted that genetic testing should be offered to patients with inherited cardiac conditions, though there are numerous challenges: access to genetic counselling; which genetic test to order; result interpretation; uncertain findings; secondary findings; and reclassification of variants over time (24,37,38). Despite this, the benefit of a genetic diagnosis for a patient and their family cannot be underestimated. Historically, we have advocated the role of genetic testing in guiding cascade testing of at-risk relatives, allowing us to determine which relatives have the family-specific variant and should continue clinical surveillance. Indeed, the option to release at-risk relatives from ongoing surveillance is the key driver making genetic testing cost-effective compared to clinical screening alone (39,40).

The role of genetic testing in clarifying diagnosis and guiding management is less well described, though studies are now increasingly supporting this approach. For example, genotype-specific evaluation of cases with pathogenic *DSP* variants allowed recognition of the phenotype as a left-dominant ACM and highlighted that the 2010 TaskForce Criteria (18) are inadequate for clinical diagnosis (36). Increasingly, anchoring access to genetic testing to poorly defined clinical classifications present a critical problem. Rather, an approach that employs key indicators of monogenic disease to better stratify individuals most likely to receive a genetic diagnosis could have greater utility. Positive family history provides high certainty of a shared genetic cause for disease and has been previously associated with a greater diagnostic yield of genetic testing for multiple inherited cardiac conditions (4,10). Phenotype extremes and severe outcomes are likewise paired with monogenic disease, indeed those with multiple LP/P or VUS in sarcomere genes are known to have more severe disease (4). Disease onset at a young age can also indicate a monogenic cause (8,9), and in those with severe disease and very young onset, *de novo* causes are often implicated (41,42). Taken together, what we understand about rare genetic variants causing disease could better inform our approach to improving the genetic yield among individuals with inherited cardiac conditions.

For those unlikely to have a strong genetic component to their disease, the contribution of many common variants to disease risk, in combination with environmental factors, is strongly suspected. Polygenic risk scores (PRS), providing a weighting of these common variants, will explain disease for a proportion of patients (5,6,43). In HCM, this is likely to be ∼40%, including those who have no family history of disease, no causative variants and overall later onset and milder phenotype compared to sarcomere-positive HCM (4,11). The overlap between monogenic and polygenic disease has been previously described, including among those with hypercholesterolemia where the highest PRS deciles, inferring the greatest risk, give rise to phenotypes equivalent to those with a monogenic basis (44). Identifying those with a complex basis for their disease will have marked clinical utility, leading to revised inheritance risks and clinical surveillance advice for relatives. Future research could examine whether risk factors for poor outcomes hold true for this group and whether precision therapies currently on the horizon will be equally as effective as those for monogenic disease. Delineating this group can mean we remove them from our “unsolved” group and better define the diagnostic yield of genetic testing.

Consideration of a complex or monogenic disease basis will allow us to better focus resource intensive gene discovery efforts. Patients who remain gene elusive following genetic testing and with monogenic disease characteristics, are enriched for new insights and discoveries that could have a transformative impact on our understanding of inherited cardiac conditions. A genetic diagnosis can be made in a further 5% of probands through additional research effort. However, the impact of applying more systematic deep phenotyping and cutting-edge genomic analysis tools to this group is currently unknown. The comprehensive, multidisciplinary and multi-omics approach by undiagnosed disease networks (45), has demonstrated the potential to discover new genes, uncover new disease mechanisms, expand phenotypes of known diseases and identify new rare diseases. Such discoveries could offer important insights into the underlying biology and potential drug targets, spurring precision management and novel therapies. Solving the cause of disease in these individuals will take bespoke and resource intensive efforts; therefore, methods to prioritise cases most likely to have an underlying monogenic disease are critical.

Our findings support genetic testing being offered to *all* patients with known or suspected heritable disease, with LP/P variants identified in 21% of those considered to have a likely complex disease basis and suspicious VUS in an additional 8%. We caution that poor diversity of genetic reference databases critically limits our ability to interpret genetic findings in underrepresented populations. Among our cohort, 15-20% of non-European background individuals had suspicious VUS identified, compared to only 10% of Europeans. Not only does this translate to fewer patients with a meaningful genetic result, allowing their family members to have cascade genetic testing, but increases the risk of a variant being misclassified with potential for harm (46). Efforts to increase diversity and be more inclusive in research studies will ensure the discoveries we make apply to the broader community.

Our study has important limitations, including that it was a retrospective audit with risk of bias in medical record review. The proposed monogenic disease scoring system was developed based on the literature and our clinical experiences, and applying it retrospectively to our cohort supported its clinical value; however, further independent validation is needed. Additionally, the cohort were those referred to a highly specialised adult clinic and may not be representative of the general population; nonetheless, the consecutive patient population makes our data applicable to “real-world” specialised clinic settings.

## Conclusion

We present an 18-year experience of consecutive patients attending a specialised cardiac genetic clinic, highlighting the role of genomics in diagnosis and management. Genetic yield is not equitable across ancestry groups. Factors indicative of monogenic disease can identify those most likely to achieve a genetic diagnosis, even when the clinical phenotype is uncertain. We propose triaging of cases with characteristics suggestive of a monogenic disease basis for additional, comprehensive research-based sequencing.

## Data Availability

All data produced in the present study are available upon reasonable request to the authors and according to ethical approval.

## Supplementary Tables

STable 1: Tier 1 and Tier 2 gene lists

STable 2: Classified variants

STable 3: Classified suspicious VUS

