## Supplementary Material for "The Role of Genetic Testing in Diagnosis and Care of Inherited Cardiac Conditions in a Specialised Multidisciplinary Clinic"

**SUPPLEMENTARY TABLE 1: Tier 1 and tier 2 gene lists**

| **Condition** | **Tier 1** | **Tier 2** |
| --- | --- | --- |
| HCM | MYBPC3 | FLNC |
|  | MYH7 | ACTN2 |
|  | TNNT2 | FHOD3 |
|  | TNNI3 | ALPK3 |
|  | ACTC1 | CSRP3 |
|  | TPM1 | JHP2 |
|  | MYL2 | PTPN11* |
|  | MYL3 | RAF1* |
|  | PRKAG2* | RIT1* |
|  | TTR* | CACNA1C* |
|  | LAMP2* | MT-T1* |
|  | GLA* | DES* |
|  |  | FHL1* |
| DCM | BAG3 |  |
|  | DES |  |
|  | FLNC (tv) |  |
|  | LMNA |  |
|  | MYH7 |  |
|  | PLN |  |
|  | RBM20 |  |
|  | SCN5A |  |
|  | TNNC1 |  |
|  | TNNT2 |  |
|  | TTN |  |
|  | DSP |  |
| Brugada syndrome | SCN5A | SCN1B |
| CPVT | RYR2 |  |
|  | CASQ2 |  |
| LQTS | KCNH2 | KCNJ2* |
|  | KCNQ1 | CALM1 |
|  | SCN5A | CALM2 |
|  | TRDN | CALM3 |
|  | KCNE1 | CACNA1C* |
|  | KCNE2 |  |
| LVNC | ACTC1 |  |
|  | ACTN2 |  |
|  | MYBPC3 |  |
|  | MYH7 |  |
|  | TAZ |  |
|  | TNNT2 |  |
|  | TPM1 |  |
|  | NKX2-5 |  |
|  | PRDM`6 |  |
| ACM | PKP2 | DES |
|  | DSC2 | FLNC |
|  | DSG2 | JUP |
|  | DSP | LDB3 |
|  | JUP | LMNA |
|  | TMEM43 | NKX2-5 |
|  |  | PLN |
|  |  | RBM20 |
|  |  | SCN5A |
| RCM | MYH7 | FLNC (missense) |
|  | TNNI3 |  |
|  | TNNT2 |  |
|  | ACTC1 |  |

***** Genes associated with common genocopies for the known phenotype

**Abbreviations:** HCM, hypertrophic cardiomyopathy; DCM, dilated cardiomyopathy; tv, truncating variant; CPVT, catecholaminergic polymorphic ventricular tachycardia; LQTS, long QT syndrome; LVNC, left-ventricular non-compaction; ACM, arrhythmogenic cardiomyopathy; RCM, restricted cardiomyopathy.

**SUPPLEMENTARY TABLE 2: Classified likely pathogenic and pathogenic variants**

| **Gene** | **Transcript** | **DNA Change** | **Protein Change** | **gnomAD V2.1.1 AC** | **gnomAD v3 AC** | **Proband Count (incl ours)** | **ACMG Criteria** | **Classification** | **Clinical Diagnosis** |
| --- | --- | --- | --- | --- | --- | --- | --- | --- | --- |
| ACTN2 | NM_001103.3 | c.355G>A | p.Ala119Thr | 0 | 0 | 3 | PP1_Strong, PM2, PP3, PS4_Supporting | Likely pathogenic | LVNC |
| CACNA1C | NM_001129827.1 | c.1553G>A | p.Arg518His | 1 | 1 | 4 | PM2, PP1_Moderate, PP3, PS4_Sup | Likely pathogenic | Undiagnosed |
| DES | NM_001927.3 | c.376G>C | p.Val126Leu | 0 | 0 | 1 | PS2, PM2, PP3 | Likely pathogenic | DCM |
| DES | NM_001927.3 | c.1360C>T | p.Arg454Trp | 0 | 0 | 16 | PS2, PS3, PS4, PM2, PP3 | Pathogenic | Possible HCM |
| DSP | NM_004415.2 | c.1755dupA | p.His586Thrfs*9 | 0 | 0 | 2 | PVS1, PM2, PP1, PP4 | Pathogenic | ARVC |
| DSP | NM_004415.2 | c.7641C>G | p.Tyr2547Ter | 0 | 1 | 1 | PVS1, PM2 | Likely pathogenic | OHCA |
| FHL1 | NM_004415.2 | c.396del | p.Cys132Ter | 0 | 0 | 1 | PVS1, PM2 | Likely pathogenic | Possible HCM |
| FHL1 | NM_001449.4 | c.596_597CGT>C | p.Arg199LeufsTer44 | 0 | 0 | 1 | PVS1_Strong, PM2 | Suspicious VUS | HCM |
| FHL1 | NM_001449.4 | c.738C>G | p.His246Gln | 0 | 0 | 2 | PM2, PP1_Moderate, PP3, PS4_Sup | Likely pathogenic | HCM |
| FHOD3 | NM_001281740.2 | c.1580_1582del | p.Ser527del | 1 | 1 | 14 | PS4, PP1_Strong, PM2 | Pathogenic | HCM |
| FLNC | NM_001127487.1 | c.4926_4927insACGTCACA | p.Val1643Thrfs*26 | 1 | 2 | 5 | PVS1, PM2, PP3 | Pathogenic | DCM |
| FLNC | NM_001127487.1 | c.617G>A | p.W206* | 0 | 0 | 1 | PVS1, PM2 | Likely pathogenic | OHCA |
| FLNC | NM_001127487.1 | c.7496_7497insTGCT | p.Gln2499Hisfs*46 | 0 | 0 | 3 | PVS1, PM2 | Pathogenic | DCM |
| LZTR | NM_001458.4 | c.406T>C | p.Tyr136His | 0 | 0 | 3 | PS2, PM2, PS4_Sup | Pathogenic | Noonan's Syndrome |
| GLA | NM_000169.2 | c.1087C>T | p.Arg363Cys | 2 | 0 | 6 | PS4, PM2, PM5, PP4 | Likely pathogenic | HCM |
| KCNH2 | NM_000238.3 | c.2453C>T | p.Ser818Leu | 1 | 0 | 7 | PP1_Strong, PM1, PM2, PM6, PS4_Moderate, PP3 | Pathogenic | LQTS |
| KCNH2 | NM_000238.3 | c.1909G>T | Glu637* | 0 | 0 | 1 | PVS1, PM1, PM2 | Pathogenic | LQTS |
| KCNH2 | NM_000238.3 | c.2044delG | p.Glu682Serfs*32 | 0 | 0 | 1 | PVS1, PM2, PP1 | Pathogenic | LQTS |
| KCNH2 | NM_000238.3 | c.1280A>G | p.Tyr427Cys | 0 | 0 | 3 | PM2, PP2, PP3, PS4_Sup | Likely Pathogenic | LQTS |
| KCNH2 | NM_000238.3 | c.1886A>G | p.Asn629Ser | 0 | 0 | 3 | PS1, PS3, PM2, PS4_Sup | Pathogenic | LQTS |
| KCNH2 | NM_000238.3 | c.3040C>T | p.Arg1014* | 0 | 1 | 5 | PVS1, PM2 | Pathogenic | LQTS |
| KCNH2 | NM_000238.3 | c.3102_3103del | p.Arg1035Glyfs*83 | 0 | 0 | 1 | PVS1, PM2 | Pathogenic | LQTS |
| KCNH2 | NM_000238.3 | c.577del | p.Ala193Profs*8 | 0 | 0 | 1 | PVS1, PM2 | Pathogenic | LQTS |
| KCNH2 | NM_000238.3 | c.1128G>A | p.Gln376= | 0 | 0 | 12 | PS4, PM2, PP3 | Likely pathogenic | LQTS |
| KCNQ1 | NM_000218.2 | c.1664G>A | Arg555His | 0 | 1 | 2 | PM2, PM5, PP3, PS4_Sup | Likely pathogenic | LQTS |
| KCNQ1 | NM_000218.2 | c.612C>G | p.Ile204Met | 0 | 0 | 1 | PM2, PM5, PS4_Sup, PS3_Sup | Likely pathogenic | LQTS |
| KCNQ1 | NM_000218.2 | c.1663C>T | p.Arg555Cys | 3 | 0 | 9 | PS3, PM2, PS4_Mod, PP1, PP3 | Pathogenic | LQTS |
| KCNQ1 | NM_000218.2 | c.477+5G>A |  | 3 | 1 | 13 | PS4, PM2 | Likely pathogenic | LQTS |
| KCNQ1 | NM_000218.2 | c.1616G>A | p.Arg539Gln | 0 | 0 | 2 | PM2, PM5, PP3, PS4_Sup | Likely pathogenic | LQTS |
| KCNQ1 | NM_000218.2 | c.1831_1834dup | p.Met612fs | 0 | 0 | 1 | PVS1, PM2 | Likely pathogenic | LQTS |
| KCNQ1 | NM_000218.2 | (1032+1_1033-1)_(1128+1_1129-1)del |  | 0 | 0 | 1 | PM2, PM4, PP4 | Likely pathogenic | LQTS |
| KCNQ1 | NM_000218.2 | c.1033-2A>G |  | 0 | 0 | 1 | PVS1, PM2 | Pathogenic | LQTS |
| KCNQ1 | NM_000218.2 | c.1022C>T | p.Ala341Val | 0 | 0 | >25 | PS3, PS4, PM2, PP1 | Pathogenic | LQTS |
| KCNQ1 | NM_000218.2 | c.944A>G | p.Tyr315Cys | 0 | 0 | 2 | PS1, PS3, PM2 | Likely pathogenic | LQTS |
| KCNQ1 | NM_000218.2 | c.828_830del | p.Ser277del |  |  |  | PVS1, PM2 | Pathogenic | LQTS |
| KCNQ1 | NM_000218.2 | c.1552C>T | p.Arg518X | 26 | 5 | 12 | PVS1, PS3, PS4, PP1, PP3 | Pathogenic | LQTS |
| KCNQ1 | NM_000218.2 | c.1022C>A | p.Ala341Glu | 0 | 0 | 14 | PS4, PS3, PM2, PM5, PP1, PP3 | Pathogenic | LQTS |
| KCNQ1 | NM_000218.2 | c.573_577delGCGCT | p.Arg192Cysfs*91 | 0 | 3 | 6 | PVS1, PM2, PS4_Mod | Pathogenic | LQTS |
| LAMP2 | NM_002294.2 | c.183+2A>G |  | 0 | 0 | 2 | PVS1, PM2 | Likely pathogenic | HCM |
| LMNA | NM_170707.3 | c.646C>T | p.Arg216Cys | 2 | 1 | 11 | PS4, PP1_Strong, PM2, PP3 | Pathogenic | HCM |
| MT-TI |  | m.4300A>G |  |  |  |  |  | Pathogenic | HCM |
| MYBPC3 | NM_000256.3 | c.1624G>C | p.Glu542Gln | 5 | 9 | >30 | PS3, PS4, PM2, PP3 | Pathogenic | HCM |
| MYBPC3 | NM_000256.3 | c.1624+4A>T |  | 3 | 4 | 18 | PVS1, PS3, PS4, PM2, PP1_Mod | Pathogenic | HCM |
| MYBPC3 | NM_000256.3 | c.3697C>T | p.Gln1233Ter | 2 | 0 | 10 | PVS1, PM2, PP3, PS4_Mod | Pathogenic | HCM |
| MYBPC3 | NM_000256.3 | c.2308+1G>A |  | 0 | 0 | 19 | PVS1, PS4, PM2 | Pathogenic | HCM |
| MYBPC3 | NM_000256.3 | c.3192dupC | p.Lys1065Glnfs*12 | 0 | 0 | 11 | PVS1, PM2, PP3, PS4_Mod, PP1 | Pathogenic | HCM |
| MYBPC3 | NM_000256.3 | c.2864_2865del | p.Pro955Argfs*95 | 0 | 1 | >20 | PVS1, PS4, PP1_Strong, PM2 | Pathogenic | HCM |
| MYBPC3 | NM_000256.3 | c.25+1G>A |  | 0 | 0 | 5 | PVS1, PM2, PP3, PS4_Sup | Pathogenic | HCM |
| MYBPC3 | NM_000256.3 | c.1504C>T | p.Arg502Trp | 13 | 14 | >50 | PS4, PP2 | Pathogenic | HCM |
| MYBPC3 | NM_000256.3 | c.3712_3713del | p.Leu1238Glyfs*3 | 0 | 0 | 1 | PVS1, PM2 | Pathogenic | HCM |
| MYBPC3 | NM_000256.3 | c.3617del | p.Gly1206Valfs*31 | 0 | 0 | 2 | PVS1, PM2, PM4, PP1, PS4_sup | Likely pathogenic | HCM |
| MYBPC3 | NM_000256.3 | c.2413+1G>A |  | 0 | 0 | 2 | PVS1, PM2, PP1, PS4_Sup | Pathogenic | HCM |
| MYBPC3 | NM_000256.3 | c.2079_2082dupCCCA | p.Ala695Profs*14 | 0 | 0 | 1 | PVS1, PM2 | Likely pathogenic | HCM |
| MYBPC3 | NM_000256.3 | c.2373dupG | p.Trp792Valfs*41 | 3 | 3 | >50 | PVS1, PS4, PP1_Strong, PM2 | Pathogenic | HCM |
| MYBPC3 | NM_000256.3 | c.2780_2781del | p.Thr927Ilefs*123 | 0 | 1 | 3 | PVS1, PM2, PS4_Sup | Pathogenic | HCM |
| MYBPC3 | NM_000256.3 | c.1351+2T>C |  | 0 | 0 | 1 | PVS1, PM2 | Likely pathogenic | HCM |
| MYBPC3 | NM_000256.3 | c.2738-2A>T |  | 0 | 0 | 2 | PVS1, PM2, PP3 | Likely pathogenic | HCM |
| MYBPC3 | NM_000256.3 | c.3624del | p.Lys1209Serfs*28 | 2 | 1 | 14 | PVS1, PM2, PS4_Mod | Pathogenic | HCM |
| MYBPC3 | NM_000256.3 | c.1928-2A>G | IVS20-2A>G | 0 | 0 | >40 | PVS1, PS4, PP1_Strong, PM2 | Pathogenic | HCM |
| MYBPC3 | NM_000256.3 | c.162delG | p.Lys54Asnfs*13 | 0 | 0 | 3 | PVS1, PM2, PS4_Sup | Pathogenic | HCM |
| MYBPC3 | NM_000256.3 | c.3627+2del |  | 0 | 0 | 2 | PVS1, PM2, PS4_Sup | Likely pathogenic | HCM |
| MYBPC3 | NM_000256.3 | c.913_914del | p.Phe305Profs*27 | 0 | 0 | >20 | PVS1, PM2, PS4 | Pathogenic | HCM |
| MYBPC3 | NM_000256.3 | c.2905C>T | p.Gln969Ter | 0 | 0 | 3 | PVS1, PM2, PS4_Sup | Pathogenic | HCM |
| MYBPC3 | NM_000256.3 | c.1302C>A | p.Tyr434* | 0 | 0 | 1 | PVS1, PM2, PP3 | Likely pathogenic | HCM |
| MYBPC3 | NM_000256.3 | c.2735del | p.Gly912Alafs*12 | 0 | 0 | 1 | PVS1, PM2 | Pathogenic | HCM |
| MYBPC3 | NM_000256.3 | c.2309-2A>G |  | 0 | 0 | 1 | PVS1, PM2 | Pathogenic | HCM |
| MYBPC3 | NM_000256.3 | c.2429G>A | p.Arg810His | 12 | 7 | >30 | PP1, PS4_sup, PS4 | Likely pathogenic | HCM |
| MYBPC3 | NM_000256.3 | c.442G>A | p.Gly148Arg | 13 | 15 | 16 | PS3, PP3, PP1 | Likely pathogenic | HCM |
| MYBPC3 | NM_000256.3 | c.2308G>A | p.Asp770Asn | 4 | 0 | 17 | PS4, PM2, PS4_Sup | Likely pathogenic | HCM |
| MYBPC3 | NM_000256.3 | c.3490+1G>A |  | 0 | 1 | 7 | PVS1, PM2, PS4_Mod | Pathogenic | HCM |
| MYBPC3 | NM_000256.3 | c.1090G>A | p.Ala364Thr | 0 | 0 | 9 | PS3, PM2, PS4_Mod, PP3 | Likely pathogenic | HCM |
| MYBPC3 | NM_000256.3 | c.1484G>A | p.Arg495Gln | 6 | 6 | 30 | PS4, PP1_Strong, PM2, PP3 | Pathogenic | HCM |
| MYBPC3 | NM_000256.3 | c.613C>T | p.Gln205* | 0 | 0 | 4 | PVS1, PM2, PS4_Sup | Pathogenic | HCM |
| MYBPC3 | NM_000256.3 | c.1153_1168del | Val385fs | 0 | 0 | 1 | PVS1, PM2 | Pathogenic | HCM |
| MYBPC3 | NM_000256.3 | c.927-9G>A | g_7301_IVS12-9_G>A | 0 | 3 | >30 | PVS1, PS4, PM2 | Pathogenic | HCM |
| MYBPC3 | NM_000256.3 | c.3476_3477insATTT | p.Phe1159Leufs*11 | 0 | 0 | 2 | PVS1, PM2, PP1, PS4_Sup | Pathogenic | HCM |
| MYBPC3 | NM_000256.3 | c.1505G>A | p.Arg502Gln | 0 | 4 | >20 | PS4, PP1_Strong, PM2, PM5 | Pathogenic | HCM |
| MYBPC3 | NM_000256.3 | c.3190+5G>A | IVS29+5 G>A | 4 | 0 | 7 | PVS1, PS3, PM2, PS4_Mod | Pathogenic | HCM |
| MYBPC3 | NM_000256.3 | c.2371C>T | p.Gln791* | 0 | 0 | 4 | PVS1, PM2, PS4_Sup | Pathogenic | HCM |
| MYBPC3 | NM_000256.3 | c.2267del | p.Pro756Leufs*66 | 0 | 0 | 1 | PVS1, PM2 | Likely pathogenic | HCM |
| MYBPC3 | NM_000256.3 | c.772G>A | p.Glu258Lys | 3 | 6 | >50 | PS3, PS4, PP1_Strong, PM2, PP3 | Pathogenic | HCM |
| MYBPC3 | NM_000256.3 | 1458-1G>A |  | 1 | 1 | 5 | PVS1, PM2, PP3, PS4_Sup | Pathogenic | HCM |
| MYBPC3 | NM_000256.3 | c.2827C>T | p.Arg943* | 2 | 4 | >25 | PVS1, PS4, PM2 | Pathogenic | HCM |
| MYBPC3 | NM_000256.3 | c.3408C>A | p.Tyr1136* | 0 | 1 | 5 | PVS1, PM2, PP1, PS4_Sup | Pathogenic | HCM |
| MYBPC3 | NM_000256.3 | c.655G>C | p.Val219Leu | 0 | 0 | 5 | PS3, PS4, PM2 | Pathogenic | HCM |
| MYBPC3 | NM_000256.3 | c.1084dup | p.Ser362LysfsTer28 | 0 | 0 | 2 | PVS1, PM2, PS4_Sup | Pathogenic | HCM |
| MYBPC3 | NM_000256.3 | c.1483C>G | p.Arg495Gly | 1 | 1 | 18 | PS4, PM2, PM5, PP3 | Pathogenic | HCM |
| MYBPC3 | NM_000256.3 | c.2558del | p.Gly853Alafs*26 | 0 | 0 | 4 | PVS1, PM2 | Likely pathogenic | HCM |
| MYBPC3 | NM_000256.3 | c.177_187delCGTGTGCCCTCT>C | p.Glu60Alafs*49 | 0 | 1 | 12 | PVS1, PS4, PM2 | Pathogenic | HCM |
| MYBPC3 | NM_000256.3 | c.2604- 2605delTCinsA | p.Ser871Alafs*8 | 0 | 0 | 12 | PVS1, PS4, PM3 | Pathogenic | HCM |
| MYBPC3 | NM_000256.3 | c.1359del | p.Val454Cysfs*12 | 0 | 0 | 1 | PVS1, PM2, PP1 | Pathogenic | HCM |
| MYBPC3 | NM_000256.3 | c.1591G>C | p.Gly531Arg | 6 | 2 | 15 | PS3, PS4, PM2, PP3 | Likely pathogenic | HCM |
| MYBPC3 | NM_000256.3 | c.821+3G>T |  | 0 | 0 | 4 | PS3, PM2, PP3 | Pathogenic | HCM |
| MYBPC3 | NM_000256.3 | c.1483C>T | p.Arg495Trp | 0 | 1 | 14 | PS4, PM2, PM5, PP3 | Likely pathogenic | HCM |
| MYBPC3 | NM_000256.3 | c.3330+2T>G |  | 0 | 5 | 9 | PVS1, PM2, PS4_mod | Pathogenic | HCM |
| MYBPC3 | NM_000256.3 | c.821+1G>C |  | 5 | 1 | >10 | PVS1, PM2, PS4_Mod | Pathogenic | HCM |
| MYBPC3 | NM_000256.3 | c.2719G>T | p.Glu907Ter | 0 | 0 | 1 | PVS1, PM2 | Pathogenic | HCM |
| MYBPC3 | NM_000256.3 | c.2096del | p.Pro699Glnfs*55 | 0 | 0 | >25 | PVS1, PM2, PS4, PP1 | Pathogenic | HCM |
| MYBPC3 | NM_000256.3 | c.3166dupG | p.Lys1055* | 0 | 0 | 7 | PVS1, PM2, PS4_Mod | Likely pathogenic | HCM |
| MYBPC3 | NM_000256.3 | c.2997del | p.Lys1000Serfs*6 | 0 | 0 | 1 | PVS1, PM2 | Likely pathogenic | HCM |
| MYBPC3 | NM_000256.3 | c.3491-3C>G |  | 0 | 0 | 1 | PVS1, PM2 | Likely pathogenic | HCM |
| MYBPC3 | NM_000256.3 | c.3163A>T | p.Lys1055* | 0 | 0 | 9 | PVS1, PM2, PS4_Mod | Pathogenic | HCM |
| MYBPC3 | NM_000256.3 | c.3190+1G>A |  | 1 | 1 | 7 | PVS1, PM2, PS4_Mod | Pathogenic | HCM |
| MYBPC3 | NM_000256.3 | c.722G>A | p.Glu258Lys | 6 | 6 | >50 | PS3, PS4, PP1_strong, PM2 | Pathogenic | HCM |
| MYBPC3 | NM_000256.3 | c.1224-52G>A |  | 1 | 3 | 3 | PS3, PM2, PS4_Sup | Likely pathogenic | HCM |
| MYBPC3 | NM_000256.3 | c.1090+453C>T |  | 0 | 0 | 1 | PS3, PM2, PP3 | Likely pathogenic | HCM |
| MYBPC3 | NM_000256.3 | c.2274C>T | p.Gly758= | 1 | 1 | 1 | PM2, PM4, PP1_Mod | Likely pathogenic | HCM |
| MYBPC3 | NM_000256.3 | c.1928-569G>T |  | 0 | 1 | 1 | PS3, PM2, PP1 | Likely pathogenic | HCM |
| MYH7 | NM_000257.2 | c.438G>T | p.Lys146Asn | 0 | 0 | 4 | PS3, PM2, PM6, PP1_Mod, PP3, PS4_Sup | Pathogenic | HCM |
| MYH7 | NM_000257.2 | c.2093T>C | p.Val698Ala | 0 | 0 | 2 | PP1_Strong, PM1, PM2, PP3, PS4_Sup | Pathogenic | HCM |
| MYH7 | NM_000257.2 | c.1816G>A | p.Val606Met | 1 | 1 | 6 | PS4, PP1_Strong, PM1, PM2, PM6, PP3 | Pathogenic | HCM |
| MYH7 | NM_000257.2 | c.1988G>A | p.Arg663His | 4 | 6 | >15 | PS3, PS4, PM2 | Pathogenic | HCM |
| MYH7 | NM_000257.2 | c.2167C>T | p.Arg723Cys | 3 | 3 | 6 | PS4, PP1_Strong, PM1, PM2, PM5 | Pathogenic | HCM |
| MYH7 | NM_000257.2 | c.2207T>C | p.Ile736Thr | 0 | 0 | >25 | PS4, PM1, PM2, PP1_Mod | Pathogenic | HCM |
| MYH7 | NM_000257.2 | c.1357C>T | p.Arg453Cys | 0 | 1 | 8 | PM1, PM2, PP1, PP3 | Pathogenic | HCM |
| MYH7 | NM_000257.2 | c.4135G>A | p.Ala1379Thr | 1 | 1 | 16 | PS4, PP1_Strong, PM2, PP3 | Pathogenic | HCM |
| MYH7 | NM_000257.2 | c.2681A>G | p.Glu894Gly | 0 | 1 | >35 | PS4, PP1_Strong, PM1, PM2, PP3 | Pathogenic | HCM |
| MYH7 | NM_000257.2 | c.1954A>G | p.Arg652Gly | 1 | 1 | 14 | PS4, PM1, PM2, PP1, PP3 | Likely pathogenic | HCM |
| MYH7 | NM_000257.2 | c.2389G>A | p.Ala797Thr | 6 | 4 | >60 | PS4, PP1_Strong, PM1, PM2 | Pathogenic | HCM |
| MYH7 | NM_000257.2 | c.2221G>T | p.Gly741Trp | 0 | 1 | >30 | PS4, PP1_Strong, PM1, PM2, PM5, PP3 | Pathogenic | HCM |
| MYH7 | NM_000257.2 | c.1207C>T | p.Arg403Trp | 0 | 0 | 12 | PS4_Mod, PM1, PM2, PP3 | Pathogenic | HCM |
| MYH7 | NM_000257.2 | c.2156G>A | p.Arg719Gln | 0 | 0 | >15 | PS4, PP1_Strong, PM1, PM2, PP3 | Pathogenic | HCM |
| MYH7 | NM_000257.2 | c.2539A>G | p.Lys847Glu | 0 | 0 | >25 | PS4, PM1, PM2, PP3 | Likely pathogenic | HCM |
| MYH7 | NM_000257.2 | c.4066G>A | p.Glu1356Lys | 0 | 1 | 24 | PS4, PM2, PP1_Mod, PP3 | Likely pathogenic | HCM |
| MYH7 | NM_000257.2 | c.715G>A | p.Asp239Asn | 1 | 0 | 12 | PS3, PP1_Strong, PM1, PM2, PS4_Mod | Likely pathogenic | HCM |
| MYH7 | NM_000257.2 | c.427C>T | p.Arg143Trp | 7 | 1 | 14 | PS4, PM2, PP1, PP3 | Likely pathogenic | HCM |
| MYH7 | NM_000257.2 | c.1208G>A | p.Arg403Gln | 0 | 0 | >30 | PS3, PS4, PP1_Strong, PM1, PM2, PM5, PP3 | Pathogenic | HCM |
| MYH7 | NM_000257.2 | c.2738T>C | p.Ile913Thr | 0 | 0 | 3 | PM1, PM2, PP3, PS4_Sup | Likely pathogenic | HCM |
| MYH7 | NM_000257.2 | c.428G>A | p.Arg143Gln | 1 | 1 | 17 | PS4, PM2, PP1, PP3 | Likely pathogenic | HCM |
| MYH7 | NM_000257.2 | c.1324C>T | p.Arg442Cys | 6 | 4 | 9 | PM1, PM2, PS4_Mod, PP1_Mod, PP3 | Pathogenic | HCM |
| MYH7 | NM_000257.2 | c.952T>G | p.Thr318Pro | 0 | 0 | 2 | PM1, PM2, PM6, PP1_Mod, PP3 | Likely pathogenic | HCM |
| MYH7 | NM_000257.2 | c.2770G>A | p.Glu924Lys | 0 | 1 | >30 | PS4, PP1_Strong, PM1, PM2, PM5, PP3 | Pathogenic | HCM |
| MYH7 | NM_000257.2 | c.2722C>G | p.Leu908Val | 1 | 2 | >15 | PS4, PM2, PM1, PP1, PP3 | Pathogenic | HCM |
| MYH7 | NM_000257.2 | c.1370T>C | p.Ile457Thr | 1 | 2 | 10 | PS4, PM1, PM2, PP3 | Likely pathogenic | HCM |
| MYH7 | NM_000257.2 | c.1727A>G | p.His576Arg | 5 | 3 | 8 | PM1, PM2, PS4_Mod, PP3 | Likely pathogenic | HCM |
| MYH7 | NM_000257.2 | c.3634C>T | p.Arg1212Trp | 0 | 0 | 1 | PM2, PP1_mod, PS4_sup | Suspicious VUS | DCM |
| MYH7 | NM_000257.2 | c.611G>A | p.Arg204His | 5 | 3 | >20 | PS4, PM1, PM2 | Likely pathogenic | HCM |
| MYH7 | NM_000257.2 | c.2539_2541del | p.Lys847del | 0 | 0 | 12 | PS4, PM2, PM4, PP1 | Likely pathogenic | HCM |
| MYH7 | NM_000257.2 | c.2609G>A | p.Arg870His | 2 | 2 | 17 | PS1, PS4, PM1, PM2, PP1, PP3 | Pathogenic | HCM |
| MYH7 | NM_000257.2 | c.4124A>G | p.Tyr1375Cys | 0 | 0 | 3 | PM2, PP1_Moderate, PP3, PS4_Sup | Likely pathogenic | HCM |
| MYH7 | NM_000257.2 | c.2155C>T | p.Arg719Trp | 1 | 1 | >30 | PS4, PP1_Strong, PM1, PM2, PM5, PP3 | Pathogenic | HCM |
| MYH7 | NM_000257.2 | c.1063G>A | p.Ala355Thr | 0 | 0 | >20 | PS4, PM1, PM2 | Likely pathogenic | HCM |
| MYH7 | NM_000257.2 | c.2146G>A | p.Gly716Arg | 0 | 0 | >20 | PS4, PP1_Strong, PM1, PM2, PM6 | Pathogenic | HCM |
| MYH7 | NM_000257.2 | c.2012G>A | p..Arg671His | 0 | 0 | 2 | PM1, PM2, PP3, PS4_Sup | Likely pathogenic | HCM |
| MYH7 | NM_000257.2 | c.5135G>A | p.Arg1712Gln | 6 | 3 | >20 | PS4, PM2, PP3 | Likely pathogenic | HCM |
| MYH7 | NM_000257.2 | c.1273G>A | p.Gly425Arg | 0 | 0 | 5 | PM1, PM2, PP2, PS4_Sup | Likely pathogenic | OHCA |
| MYH7 | NM_000257.2 | c.2333A>T | p.Asp778Val | 0 | 1 | 4 | PM1, PM2, PM5, PP3, PS4_Sup | Likely pathogenic | HCM |
| MYL2 | NM_000432.3 | c.64G>A | p.Glu22Lys | 5 | 3 | 28 | PS3, PS4, PP1_Strong, PM2 | Suspicious VUS | HCM |
| MYL3 | NM_000258.2 | c.463C>G | p.His155Asp | 0 | 0 | 3 | PP1_Strong, PM2, PP3, PS4_Supporting | Likely pathogenic | HCM |
| NDUFB11 | NM_019056.6 | c.391G>A | p.Glu131Lys | 0 | 0 | 5 | PM2, PM6, PP3, PS4_Sup | Likely pathogenic | Undiagnosed |
| NKX2-5 | NM_004387.3 | c.744C>A | p.Tyr248* | 0 | 0 | 1 | PVS1, PM1, PM2 | Pathogenic | LVNC |
| PKP2 | NM_004572.3 | c.2203C>T | p.Arg735* | 1 | 0 | 8 | PVS1, PM2, PS4_Mod, PP1 | Pathogenic | ARVC |
| PKP2 | NM_004572.3 | c.2197_2022delinsG | p.A733fs*740 | 0 | 0 | >30 | PVS1, PS4, PM2 | Pathogenic | ARVC |
| PKP2 | NM_004572.3 | c.2146-1G>C | IVS10-1G>C | 9 | 4 | >50 | PVS1, PS4, PM2, PP1 | Pathogenic | ARVC |
| PKP2 | NM_004572.3 | c.1237C>T | p.Arg413Ter | 4 | 7 | 14 | PVS1, PS3, PS4_Mod, PM2, PP1 | Pathogenic | ARVC |
| PKP2 | NM_004572.3 | c.2489+1G>A |  | 8 | 6 | >20 | PVS1, PS4, PM2 | Pathogenic | OHCA |
| PKP2 | NM_004572.3 | c.2229dupT | Leu744Serfs*3 | 0 | 0 | 1 | PVS1, PM2 | Likely pathogenic | OHCA |
| PRDM16 | NM_022114.3 | c.564delC | p.Ser189Valfs*22 | 0 | 0 | 1 | PVS1_Strong, PM2 | Likely pathogenic | LVNC |
| PRKAG2 | NM_016203.3 | c.905A>G | p.Arg302Gln | 0 | 1 | 13 | PS3, PS4, PP1_Strong, PM2 | Likely pathogenic | HCM |
| PRKAG2 | NM_016203.3 | c.773G>C | p.Arg258Pro | 0 | 0 | 1 | PM2, PS4_supp, PP1, PP3 - LP | Likely pathogenic | HCM |
| PTPN11 | NM_002834.3 | c.836A>G | p.Tyr279Cys | 0 | 0 | 1 | PM2, PP3 | Suspicious VUS | Noonan's Syndrome |
| PTPN11 | NM_002834.3 | c.1403C>T | p.Thr468Met | 1 | 1 | >50 | PS3, PS4, PM6, PP2, PP3 | Pathogenic | Noonan's Syndrome |
| RAF1 | NM_002880.3 | c.779C>T | p.Thr260Ile | 0 | 0 | 3 | PM1, PM2, PM5, PP3, PS4_Sup | Likely pathogenic | Noonan's Syndrome |
| RBM20 | NM_001134363.1 | c.3267delC | p.Ile1090Serfs*14 | 2 | 1 | 1 | PVS1, PM2 | Suspicious VUS | DCM |
| RBM20 | NM_001134363.1 | c.1904C>A | p.Ser635Tyr | 0 | 0 | 1 | PM1, PM2, PM5, PP3 | Likely pathogenic | DCM |
| RIT1 | NM_006912.5 | c.157T>C | p.Tyr53His | 0 | 0 | 3 | PS3, PS4, PM2, PP3 | Likely pathogenic | Noonan's Syndrome |
| RYR2 | NM_001035.2 | c.12272C>T | p.Ala4091Val | 0 | 0 | 3 | PM1, PM2, PM6, PP3, PS4_Sup | Pathogenic | CPVT |
| RYR2 | NM_001035.2 | c.7175A>G | p.Tyr2392Cys | 1 | 0 | 5 | PM2, PS4_Mod, PP1, PP2, PP3 | Pathogenic | CPVT |
| RYR2 | NM_001035.2 | c.12284G>A | p.Gly4095Asp | 0 | 0 | 1 | PM1, PM2, PM6, PP2, PP3 | Likely pathogenic | CPVT |
| RYR2 | NM_001035.2 | c.11217G>A | p.Met3739Ile | 0 | 0 | 1 | PM2, PM6, PP2, PP3 | Likely pathogenic | CPVT |
| SCN5A | NM_001099404.1 | c.3823G>A | p.Asp1275Asn | 2 | 0 | 9 | PP1_Strong, PM2, PS4_Mod, PP3 | Pathogenic | LQTS |
| SCN5A | NM_001099404.1 | c.2254G>A | p.G752R | 1 | 0 | 13 | PP1_Moderate, PM2, PS4_Mod, PP3 | Likely pathogenic | BrS |
| SCN5A | NM_001099404.1 | c.4086delG | p.Arg1362fs | 0 | 0 | 1 | PVS1, PM2 | Likely pathogenic | BrS |
| SCN5A | NM_001099404.1 | c.2582_2583delTT | c.2582_2583del | 1 | 0 | 20 | PVS1, PS1, PS4, PM2, PP1 | Pathogenic | BrS |
| SCN5A | NM_001099404.1 | c.1936delC | p.Gln646Argfs*5 | 0 | 2 | 8 | PVS1, PP1_strong, PM2 | Pathogenic | BrS |
| SCN5A | NM_001099404.1 | c.4772G>A | p.Trp1591* | 0 | 0 | 2 | PVS1, PM2, PS4_Sup | Pathogenic | BrS |
| SCN5A | NM_001099404.1 | c.4437 +5G>A |  | 0 | 0 | 4 | PVS1, PS3, PM2, PS4_Sup | Likely pathogenic | BrS |
| SCN5A | NM_001099404.1 | c.5228G>A | p.Gly1743Glu | 0 | 0 | 3 | PS3, PM2, PS4_Sup, PP3 | Pathogenic | BrS |
| SCN5A | NM_001099404.1 | c.4925G>A | p.Gly1642Glu | 0 | 0 | 5 | PM2, PS4_Sup, PP1, PP3 | Likely pathogenic | BrS |
| SCN5A | NM_001099404.1 | c.5126C>T | p.Thr1709Met | 1 | 0 | 11 | PM2, PS4_Mod, PP2 | Likely pathogenic | BrS |
| SCN5A | NM_001099404.1 | c.1066C>T | p.Asp356Asn | 1 | 1 | 13 | PS4, PM2, PP3 | Likely pathogenic | OHCA |
| SCN5A | NM_001099404.1 | c.2865_2866delGA | p.Glu955Aspfs*74 | 4 | 0 | 5 | PVS1, PM2 | Likely pathogenic | LQTS |
| SCN5A | NM_001099404.1 | c.5350G>A | p.Glu1784Lys | 0 | 2 | >15 | PS3, PS4, PM2, PP1, PP3 | Pathogenic | LQTS |
| SCN5A | NM_000335.4 |  | Translocation |  |  |  |  | Suspicious VUS | Possible HCM |
| SOS2 | NM_006939.2 | c.791C>A | p.Thr264Lys | 0 | 0 | 2 | PS2, PS4_Sup, PM2, PP3 | Likely pathogenic | Noonan's Syndrome |
| TBX5 | NM_000192.3 | c.105dupC | p.Ser36Thrfs*25 | 0 | 0 | 1 | PVS1, PM2 | Pathogenic | LVNC |
| TMEM43 | NM_024334.2 | c.1073C>T | p.Ser358Leu | 0 | 0 | >15 | PS2, PS4, PM2, PP1, PP3 | Pathogenic | HCM |
| TNNI3 | NM_000363.4 | c.485G>C | p.Arg162Pro | 10 | 1 | >25 | PS4, PP1_Strong, PM2 | Pathogenic | HCM |
| TNNI3 | NM_000363.4 | c.509G>A | p.Arg170Gln | 0 | 0 | 8 | PS2, PM2, PS4_Mod, PP3 | Pathogenic | HCM |
| TNNI3 | NM_000363.4 | c.433C>T | p.Arg145Trp | 3 | 3 | >30 | PS3, PS4, PP1_Strong, PM2, PP3 | Pathogenic | HCM |
| TNNI3 | NM_000363.4 | c.434G>A | p.Arg145Gln | 4 | 0 | 13 | PS4, PM2, PM5, PP3 | Likely pathogenic | HCM |
| TNNI3 | NM_000363.4 | c.370G>C | p.Glu124Gln | 2 | 0 | 7 | PM2, PP3, PS4_Mod, PM1_Sup | Suspicious VUS | HCM |
| TNNT2 | NM_001001430.2 | c.311C>T | p.Ala104Val | 4 | 0 | 9 | PM2, PP3, PS4_Mod, PM1_Sup | Suspicious VUS | HCM |
| TNNT2 | NM_001001430.1 | c.487_489del | p.Glu163del | 1 | 0 | >15 | PS4, PP1_Strong, PM2 | Pathogenic | HCM |
| TNNT2 | NM_001001430.1 | c.274C>T | p.Arg92Trp | 2 | 0 | >15 | PS1, PS3, PS4, PP1_Strong, PM2, PP3 | Pathogenic | HCM |
| TNNT2 | NM_001001430.1 | c.833G>C | p.Arg278Pro | 0 | 0 | 13 | PM2, PS4_Mod, PP3 | Likely pathogenic | HCM |
| TNNT2 | NM_001001430.1 | c.236T>A | p.Ile79Asn | 1 | 0 | >15 | PS3, PS4, PP1_Strong, PM2, PP3 | Pathogenic | HCM |
| TNNT2 | NM_001001430.1 | c.275G>A | Arg92Gln | 0 | 1 | >15 | PS3, PS4, PP1_Strong, PM2, PM6, PP3 | Pathogenic | HCM |
| TNNT2 | NM_001001430.1 | c.517_519del | p.Glu173Del | 1 | 0 | >15 | PS4, PP1_Strong, PM2 | Likely pathogenic | HCM |
| TNNT2 | NM_001276347.1 | c.330T>G | p.Phe110Leu | 0 | 0 | 10 | PS4, PM2, PP3 | Likely pathogenic | HCM |
| TNNT2 | NM_001001430.2 | c.2870C>G | Arg141Trp | 0 | 0 | 5 | PS3, PS4, PP1_Strong, PM2, PM6, PP3 | Pathogenic | DCM |
| TNNT2 | NM_000364.3 | c.613C>T | p.Arg205Trp | 0 | 0 | 3 | PS3, PM2, PS4_Sup, PP3 | Likely pathogenic | DCM |
| TPM1 | NM_001018005.1 | c.548C>T | p.Ala183Val | 4 | 2 | 12 | PS4, PM2, PP3 | Likely pathogenic | HCM |
| TPM1 | NM_001018005.1 | c.574G>A | p.Glu192Lys | 0 | 0 | >40 | PS4, PP1_Strong, PM2, PP3, PP2 | Pathogenic | HCM |
| TTN | NM_001267550.1 | c.66081_66088del | p.Gln22027Hisfs*4 | 0 | 0 | 1 | PVS1, PM2 | Likely pathogenic | DCM |
| TTN | NM_001267550.1 | c.12742C>T | p.Gln4248Ter | 1 | 0 | 1 | PM2, PVS1_Mod, PS4_Mod | Pathogenic | Undiagnosed |
| TTN | NM_001256850.1 | c.63302-1G>C |  | 0 | 0 | 1 | PVS1, PM2 | Suspicious VUS | DCM |
| TTN | NM_001256850.1 | c.59352del | p.Glu19785SerfsTer2 | 0 | 0 | 1 | PVS1, PM3 | Suspicious VUS | DCM |
| TTN | NM_001256850.1 | c.60619del | p.Tyr20207Ilefs*5 | 0 | 0 | 1 | PVS1, PM4 | Suspicious VUS | DCM |

**Abbreviations:** AC, aggregate count; ACMG, American College of Medical Genetics; LVNC, left-ventricular non-compaction; DCM, dilated cardiomyopathy; HCM, hypertrophic cardiomyopathy; ARVC, arrhythmogenic right-ventricular cardiomyopathy; OHCA, out-of-hospital cardiac arrest; LQTS, long QT syndrome; CPVT, catecholaminergic polymorphic ventricular tachycardia; BrS, Brugada syndrome.

**SUPPLEMENTARY TABLE 3: Classified suspicious variants of unknown significance**

| **Gene** | **Transcript** | **DNA Change** | **Protein Change** | **gnomAD V2.1.1 AC** | **gnomAD v3 AC** | **Proband Count (incl ours)** | **ACMG Criteria** | **Classification** | **Clinical Diagnosis** |
| --- | --- | --- | --- | --- | --- | --- | --- | --- | --- |
| ACTC1 | NM_005159.4 | c.850A>T | p.Ile284Phe | 0 | 0 | 1 | PM2, PP3 | Suspicious VUS | HCM |
| ACTC1 | NM_005159.5 | c.499A>G | p.Ile167Val | 0 | 0 | 1 | PM2 | Suspicious VUS | HCM |
| ALPK3 | NM_020778.5 | c.496_509>del | p.Val166Hisfs*4 | 0 | 1 | 1 | PVS1_mod, PM2 | Suspicious VUS | HCM |
| DSG2 | NM_001943.3 | c.961T>A | p.Phe321Ile | 0 | 0 | 1 | PM2 | Suspicious VUS | ARVC |
| FHOD3 | NM_001281740.2 | c.1571_1579dup | p.Phe526_Ser527insTyrLeuPhe | 1 | 0 | 1 | PM2 | Suspicious VUS | HCM |
| FHOD3 | NM_001281740.3 | c.1646+1G>C |  | 0 | 0 | 2 | PM2, PP3 | Suspicious VUS | HCM |
| FLNC | NM_001458.4 | c.5692G>A | p.Gly1898Ser | 0 | 0 | 1 | PM2, PP3 | Suspicious VUS | HCM |
| FLNC | NM_001458.5 | c.7462G>A | p.Gly2488Ser | 2 | 0 | 1 | PM2, PP3 | Suspicious VUS | HCM |
| FLNC | NM_001458.6 | c.2818A>G | p.Met940Val | 0 | 0 | 1 | PM2 | Suspicious VUS | HCM |
| FLNC | NM_001458.7 | c.4871C>T | p.Ser1624Leu | 1 | 0 | 2 | PM2, PP3, PP1, PS3_Sup | Suspicious VUS | HCM |
| GLA | NM_000169.2 | c.644A>G | p.Asn215Ser | 1 | 0 | 1 | PM2 | Suspicious VUS | HCM |
| KCNE2 | NM_172201.1 | c.362T>A | p.Met121Lys | 2 | 1 | 2 | PM2 | Suspicious VUS | LQTS |
| KCNH2 | NM_000238.3 | c.1277C>T | p.Pro426Leu | 0 | 0 | 2 | PM2, PM6, PP2, PP3 | Likely pathogenic | LQTS |
| KCNH2 | NM_000238.3 | c.2345T>C | p.Ile782Thr | 0 | 0 | 1 | PM2; PP3; PP1_sup | Suspicious VUS | LQTS |
| KCNH2 | NM_000238.3 | c.281T>C | p.Val94Ala | 0 | 0 | 1 | PM2, PP3 | Suspicious VUS | LQTS |
| KCNH2 | NM_000238.3 | c.2588G>A | p.Arg863Gln | 0 | 0 | 5 | PM2, PP3, PP2 | Suspicious VUS | LQTS |
| MYBPC3 | NM_000256.3 | c.2234A>G | p.Asp745Gly | 0 | 0 | 4 | PM2, PP3, PS4_sup | Suspicious VUS | HCM |
| MYBPC3 | NM_000256.3 | c.3700G>A | p.Gly1234Arg | 0 | 0 | 1 | PM2, PP3 | Suspicious VUS | HCM |
| MYBPC3 | NM_000256.3 | c.654+5G>C |  | 0 | 0 | 2 | PM2, PP3, PS4_sup | Suspicious VUS | HCM |
| MYBPC3 | NM_000256.3 | c.3065G>A | p.Arg1022His | 8 | 0 | 1 | PM2, PP3 | Suspicious VUS | HCM |
| MYBPC3 | NM_000256.3 | c.3642G>T | p.Trp1214Cys | 0 | 0 | 1 | PM2, PP3 | Suspicious VUS | HCM |
| MYBPC3 | NM_000256.3 | c.1429G>A | p.Val477Ile | 0 | 1 | 1 | PM2, PP3 | Suspicious VUS | HCM |
| MYBPC3 | NM_000256.3 | c.3728C>G | p.Pro1243Arg | 0 | 0 | 2 | PM2, PS4_sup, PP3 | Suspicious VUS | HCM |
| MYBPC3 | NM_000256.3 | c.2449C>G | p.Arg817Gly | 0 | 0 | 3 | PM2, PP3, PS4_sup | Suspicious VUS | HCM |
| MYBPC3 | NM_000256.3 | c.3277G>T | p.Gly1093Cys | 9 | 2 | 6 | PP3, PM2, PS4_Mod | Suspicious VUS | HCM |
| MYBPC3 | NM_000256.3 | 3751T>C | Tyr1251His | 0 | 0 | 7 | PM2, PP3, PS4_Mod | Suspicious VUS | HCM |
| MYBPC3 | NM_000256.3 | c.1765G>A | p.Arg589Cys | 2 | 0 | 5 | PM2, PP3, PS4_Sup | Suspicious VUS | HCM |
| MYBPC3 | NM_000256.3 | c.3599T>C | p.Leu1200Pro | 0 | 0 | 2 | PM2, PP3 | Suspicious VUS | HCM |
| MYBPC3 | NM_000256.3 | c.3343G>A | p.Val1115Ile | 3 | 4 | 4 | PM2, PS4_sup, PP3 | Suspicious VUS | HCM |
| MYBPC3 | NM_000256.3 | c.3548T>G | p.Phe1183Cys | 0 | 1 | 2 | PM2, PP3 | Suspicious VUS | HCM |
| MYBPC3 | NM_000256.3 | c.3798C>G | p.Cys1266Trp | 0 | 0 | 7 | PM2, PP3, PS4_Mod | Suspicious VUS | HCM |
| MYBPC3 | NM_000256.3 | c.3751T>C | p.Tyr1251His | 0 | 0 | 7 | PM2, PP3, PS4_Mod | Suspicious VUS | HCM |
| MYBPC3 | NM_000256.3 | c.2543C>A | p.Ala848Glu | 0 | 0 | 4 | PM2, PP1, PP3, PS4_Sup | Suspicious VUS | HCM |
| MYH7 | NM_000257.2 | c.2225C>A | p.Ala742Glu | 0 | 0 | 1 | PM1, PM2, PP3 | Suspicious VUS | HCM |
| MYH7 | NM_000257.2 | c.3170G>A | p.Gly1057Asp | 0 | 0 | 4 | PM2, PP3, PS4_Sup | Suspicious VUS | HCM |
| MYH7 | NM_000257.2 | c.5533C>T | p.Arg1845Trp | 0 | 0 | 1 | PM2, PP3 | Suspicious VUS | Possible HCM |
| MYH7 | NM_000257.2 | c.556G>C | p.Val186Leu | 0 | 0 | 2 | PM1, PM2, PS4_Sup | Suspicious VUS | HCM |
| MYH7 | NM_000257.2 | c.4954G>T | p.Asp1652Tyr | 12 | 1 | 4 | PM2, PP1, PP3 | Suspicious VUS | HCM |
| MYH7 | NM_000257.2 | c.3325A>G | p.Lys1109Glu | 0 | 0 | 1 | PM2, PP2, PP3 | Likely pathogenic | HCM |
| MYH7 | NM_000257.2 | c.3064A>G | p.Lys1022Glu | 0 | 0 | 1 | PM2, PP3 | Suspicious VUS | HCM |
| MYH7 | NM_000257.2 | c.5134C>T | p.Arg1712Trp | 0 | 1 | 3 | PM2, PP3, PS4_sup | Suspicious VUS | HCM |
| MYH7 | NM_000257.2 | c.3592G>T | p.Asp1198Tyr | 0 | 0 | 1 | PM2, PP3 | Suspicious VUS | HCM |
| MYH7 | NM_000257.2 | c.2578A>G | p.Lys860Glu | 0 | 1 | 1 | PM1, PM2, PP3 | Suspicious VUS | HCM |
| MYH7 | NM_000257.2 | c.2585C>T | p.Ala862Val | 8 | 6 | 3 | PM2, PP3, PS4_Sup | Suspicious VUS | HCM |
| MYH7 | NM_000257.2 | c.3523C>T | p.Arg1175Trp | 5 | 4 | 1 | PM2, PP3 | Suspicious VUS | HCM |
| MYH7 | NM_000257.2 | c.4075C>T | p.Arg1359Cys | 2 | 1 | 5 | PM2, PS4_Sup, PP3 | Suspicious VUS | DCM |
| MYH7 | NM_000257.2 | c.532G>A | p.Gly178Arg | 2 | 0 | 4 | PM2, PP3, PS4_Sup | Suspicious VUS | LVNC |
| MYH7 | NM_000257.2 | c.5690G>A | p.Arg1897His | 1 | 0 | 4 | PM2, PP3, PS4_Mod | Suspicious VUS | LVNC |
| MYL3 | NM_000258.2 | c.152T>C | p.Ile51Thr | 7 | 4 | 1 | PM2 | Suspicious VUS | HCM |
| MYL3 | NM_000258.2 | c.518T>C | p.Met173Thr | 0 | 0 | 1 | PM2, PP3 | Suspicious VUS | HCM |
| PKP2 | NM_004572.3 | c.1163G>A | p.Arg388Gln | 3 | 2 | 1 | PM2, PP3 | Suspicious VUS | ARVC |
| PRKAG2 | NM_016203.3 | c.1508A>G | p.Gln503Arg | 3 | 0 | 1 | PM2, PP3 | Suspicious VUS | HCM |
| PRKAG2 | NM_016203.3 | c.1315A>G | p.Ile439Val | 3 | 1 | 2 | PM2 | Suspicious VUS | HCM |
| PRKAG2 | NM_016203.3 | c.4331G>A | p.Gly145Arg | 2 | 3 | 3 | PM2, PP3 | Suspicious VUS | HCM |
| RBM20 | NM_001134363.1 | c.1699G>A | p.Ala567Thr | 0 | 0 | 1 | PM2, PP3 | Suspicious VUS | DCM |
| RYR2 | NM_001035.2 | c.14122T>G | p.Trp4708Gly | 0 | 0 | 1 | PM2, PP3 | Suspicious VUS | CPVT |
| RYR2 | NM_001035.2 | c.11146G>A | p.Glu3716Lys | 0 | 0 | 1 | PM2, PP3 | Suspicious VUS | Possible CPVT |
| SCN5A | NM_000335.4 | c.2182G>A | p.Val728Ile | 3 | 3 | 2 | PM2, PP3 | Suspicious VUS | OHCA |
| TBX5 | NM_000192.3 | c.510+5G>A |  | 0 | 0 | 1 | PM2, PP3 | Suspicious VUS | LVNC |
| TNNI3 | NM_000363.4 | c.550-1C>T |  | 0 | 0 | 1 | PM2 | Suspicious VUS | HCM |
| TNNI3 | NM_000363.4 | c. 440T>C | p. Val147Ala | 0 | 0 | 1 | PM2, PP3, PM1 | Suspicious VUS | HCM |
| TNNI3 | NM_000363.4 | c.298C>T | p.Leu100Phe | 3 | 0 | 2 | PM2, PP3, PS4_Sup | Suspicious VUS | HCM |
| TNNI3 | NM_000363.4 | c.626A>C | p.Glu209Ala | 0 | 0 | 1 | PM2, PP3, PM1 | Suspicious VUS | HCM |
| TNNI3 | NM_000363.4 | c.500T>C | p.Leu167Pro | 1 | 0 | 2 | PM2, PP3, PS4_Sup | Suspicious VUS | DCM |
| TNNI3 | NM_000363.4 | c.538G>A | p.Asp180Asn | 0 | 0 | ? | PM2, PP3, PS4_Sup | Suspicious VUS | DCM |
| TNNT2 | NM_001001430.2 | c.652G>T | p.Val218Leu | 3 | 0 | 2 | PM2, PP3, PS4_Sup | Suspicious VUS | HCM |
| TNNT2 | NM_001001430.2 | c.833G>A | p.Arg278His | 4 | 7 | 4 | PP3, PS4_Sup | Suspicious VUS | HCM |
| TNNT2 | NM_001001430.2 | c.450G>T | p.Asn150Lys | 0 | 0 | 1 | PM2, PM1_Sup | Suspicious VUS | HCM |
| TNNT2 | NM_001001430.2 | c.844A>G | p.Lys282Glu | 0 | 0 | 1 | PM2, PP3 | Suspicious VUS | HCM |
| TNNT2 | NM_001001430.2 | c.492C>A | p.Asn164Lys | 0 | 0 | 1 | PM2, PM1_Sup | Suspicious VUS | HCM |
| TPM1 | NM_001018005.1 | c.635A>T | p.Glu212Val | 1 | 0 | 1 | PM2, PP3 | Suspicious VUS | HCM |
| TPM1 | NM_001018005.1 | c.677A>G | p.Lys226Arg | 0 | 0 | 1 | PM2, PP3 | Suspicious VUS | HCM |

**Abbreviations:** AC, aggregate count; ACMG, American College of Medical Genetics; VUS, variant of unknown significance; HCM, hypertrophic cardiomyopathy; ARVC, arrhythmogenic right ventricular cardiomyopathy; LQTS, long QT syndrome; DCM, dilated cardiomyopathy; LVNC, left-ventricular non-compaction; CPVT, catecholaminergic polymorphic ventricular tachycardia; OHCA, out-of-hospital cardiac arrest.
